# Cross-database validation reveals distinct layers of transportability in ICU delirium prediction

**DOI:** 10.64898/2026.07.19.26358409

**Authors:** Shengliang Ni, Kengo Sato

**Affiliations:** School of Life Science and Technology, Institute of Science Tokyo, 2-12-1-M6-12, Ookayama, Meguro-ku, 152-8550, Tokyo, Japan

**Author notes:** Contributing authors.

**Keywords:** ICU delirium, transportability, electronic health records, external validation, model calibration, clinical prediction

## Abstract

External validation of clinical AI emphasizes discrimination, although deployment requires the endpoint, probability estimates and operating policy to transport. Here we show that these layers diverged in retrospective bidirectional evaluation of five model families across eICU and MIMIC-IV. Coarse-label AUROC fell from 0.87–0.92 internally to 0.66–0.83 during source-only transfer. For assessment-conditioned repeated monitoring of persistence or recurrence, external AUROC reached 0.76–0.94, but removing assessment history reduced it by 0.16–0.32; broader features did not help consistently. Transported scores concentrated future-positive ICU stays 2.4–6.9-fold in the top risk decile. Development-selected cutoffs alerted 0.3–2.0% of prediction rows and captured 9.2–11.0% of future-positive rows; after deduplication, 4.9–12.2% of stays were alerted, capturing 43.9–49.4% of future-positive stays. Thus, ranking can persist while probability and policy transport remain site dependent. Layered validation is a prerequisite for prospective evaluation, not evidence of clinical benefit.

## 1 Introduction

Delirium in critically ill patients is a fluctuating acute disturbance of attention, awareness and cognition whose recognition depends on repeated bedside assessment [1, 2]. Unlike continuously monitored physiological signals, delirium is observed intermittently through structured instruments such as the Confusion Assessment Method for the ICU (CAM-ICU) and Intensive Care Delirium Screening Checklist (ICDSC) [3, 4]. Assessment timing, inability to assess during deep sedation, local documentation and clinical workflow therefore determine which states enter the electronic health record (EHR) and when. Recent EHR studies likewise report infrequent or discontinuous CAM/CAM-ICU measurements and construct prediction cohorts around available assessments [5–7]. A model can consequently learn both the patient’s recorded state and the process by which that state was sought and documented. ICU delirium prediction is therefore a label-observation and transportability problem, not only a model-selection problem.

Delirium prediction has progressed from admission-time risk scores to nonlinear EHR models and repeated short-horizon forecasts. PRE-DELIRIC estimated whole-stay risk from early clinical factors and was subsequently recalibrated internationally, whereas E-PRE-DELIRIC used admission-time predictors across ICUs in several countries [8–10]. Later studies evaluated random forests and gradient-boosted trees on broader clinical representations in inpatient, perioperative and critical-care cohorts [5, 11–14]. ICU studies introduced repeated-anchor, short-horizon and recurrent formulations [6, 7, 15], and recent postoperative delirium studies modelled intraoperative time series or raw electroencephalography with deep neural networks [16, 17]. These approaches expanded modelling capability, but they were developed around different prediction times, outcome rules and assessment-conditioned populations. Their reported performance therefore does not describe one interchangeable clinical task [18, 19].

Clinical portability therefore requires more than preserved discrimination. The endpoint and eligible population must correspond to the intended decision; score ordering must remain informative; predicted probabilities must retain meaning; and thresholds must yield an acceptable balance of workload and review yield [20–22]. Existing external evidence has exposed individual parts of this chain. The PRIDE study observed substantial attenuation when an admission-time ICU model was evaluated in MIMIC-III, and recent single-centre postoperative models identify external validation as an unmet requirement [16, 17, 23]. A multimodal general-ward model was associated with greater delirium detection and changes in medication use during live implementation, demonstrating feasibility, although its retrospective pre–post analysis did not establish causal clinical benefit or ICU transportability [24]. What remains unresolved is whether endpoint validity, risk ranking, probability calibration and review policy fail together or separately when a dynamic ICU model crosses data systems.

Here we treated transportability as a layered empirical question rather than a single performance value. Across the eICU Collaborative Research Database (eICU) and Medical Information Mart for Intensive Care IV (MIMIC-IV) [25, 26], we compared carried-forward and direct-assessment operational endpoints, trained five model families and performed internal and bidirectional source-only external validation across temporal settings. We then used feature expansion, input attribution and history ablation to identify what sustained external ranking, and used supervised target-label adaptation, risk concentration, calibration, decision curves and development-selected frozen two-cutoff analyses to test whether transported scores retained probability and policy meaning. All endpoints were operational sensitivity targets rather than adjudicated diagnoses. This design separates internal learnability from endpoint, ranking, probability and policy transport, while limiting clinical conclusions to retrospective risk stratification, supervised local updating and clinical-review burden rather than prospective actionability or intervention benefit.

## 2 Results

### 2.1 Database-specific observation processes defined different prediction populations

Before comparing models, we tested whether eICU and MIMIC-IV represented the same delirium-prediction setting. We mapped ICU stays from the multi-centre eICU database and the single-centre MIMIC-IV database onto a common temporal grid, with eligible direct-assessment anchors beginning after a 24 h history period (Fig. 1a and Supplementary Fig. S23). The resulting cohorts comprised 2,496,432 windows from 28,321 eICU ICU stays and 5,881,343 windows from 49,828 MIMIC-IV ICU stays. All subsequent analyses used the same sequence of label harmonization, feature construction, model development and bidirectional cross-database validation.

**Fig. 1.**
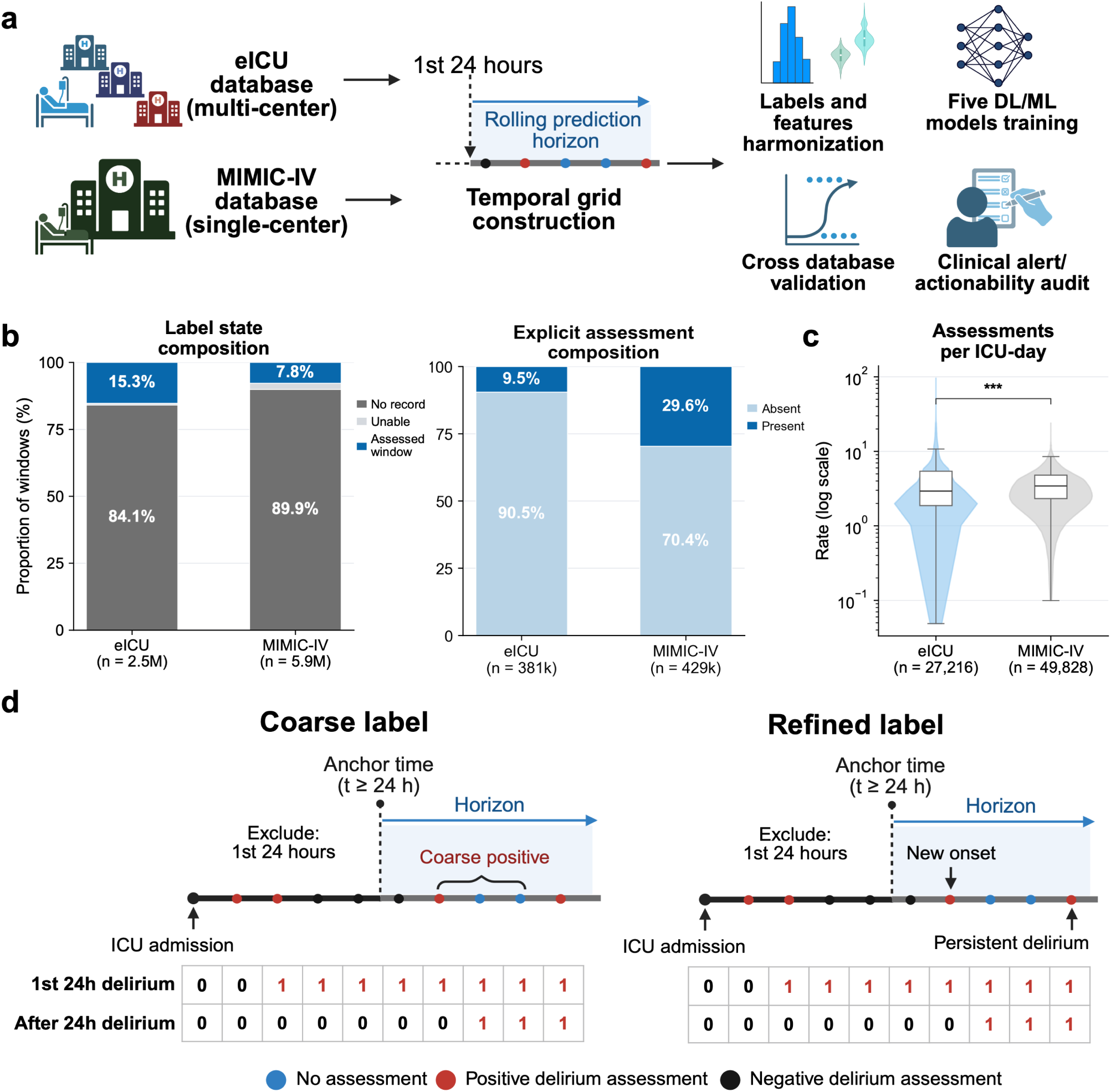
Database-specific delirium observation and operational endpoint construction. **a**, eICU and MIMIC-IV stays were mapped to a common temporal grid after a 24 h history period, followed by label and feature harmonization, training of five machine-learning or deep-learning model families, bidirectional cross-database validation and retrospective clinical-review and alert-burden analyses. Colored points on the timeline denote explicit positive, explicit negative and unassessed times; the arrow denotes a rolling prediction horizon rather than fixed 4 h spacing between observations. The rightmost icon is schematic: no prospective actionability, intervention effectiveness, workflow benefit or patient benefit was evaluated. **b**, Window-level label-state composition (left) and explicit-assessment composition (right). Counts below bars denote the corresponding numbers of windows. **c**, Distribution of delirium assessments per ICU-day among ICU stays with assessment data; violins show distributions and embedded box plots show medians and interquartile ranges, with whiskers extending to the plotted box-plot limits. *n* denotes ICU stays. ***, *P <* 0.001, two-sided, tie- and continuity-corrected asymptotic Mann–Whitney *U* test. **d**, Schematic distinction between carried-forward and direct-assessment operational label rules. The example contrasts a coarse-positive horizon with a future new-onset event; the primary operational target in later figures was persistent/recurrent delirium. The schematic illustrates study design and does not establish that either rule is a more valid representation of clinical delirium.

The observation process differed markedly between databases (Fig. 1b,c and Supplementary Table S9). An assessed label state was available in 15.3% of eICU windows and 7.8% of MIMIC-IV windows; 84.1% and 89.9%, respectively, had no assessment record, while 0.7% and 2.3% were classified as unable to assess. Among windows with an unambiguous explicit positive or negative assessment, the positive fraction was 9.5% in eICU and 29.6% in MIMIC-IV. Assessment rates per ICU-day also differed (two-sided, tie- and continuity-corrected asymptotic Mann–Whitney *U* test, *P* = 3.12 × 10^−48^). Thus, the databases differed both in when delirium was observed and in the case mix among observed assessments.

### 2.2 Operational endpoints selected different clinical questions and cohorts

We next separated a carried-forward coarse weak-label benchmark from direct-assessment strict new-onset and persistent/recurrent targets (Fig. 1d). The coarse rule asked whether a carried-positive state appeared within the prediction horizon. The direct-assessment rules retained explicit positive, explicit negative, conflict, unable-to-assess and no-assessment states and required direct future evidence. Consequently, refined performance estimated short-horizon risk conditional on documented assessment at the anchor and again within the horizon; it did not estimate delirium risk among all ICU stays or all time windows. These rules selected different clinical questions and analysis populations; their comparison defined the scope of model performance rather than validating one rule as a more accurate representation of delirium.

The selected windows differed substantially, as expected from the endpoint definitions (Fig. 3a,b). MIMIC-IV contained 1,703,313 coarse-positive windows and eICU contained 283,436. Of these, 120,623 MIMIC-IV windows (7.1%) and 32,636 eICU windows (11.5%) had a same-window explicit-positive assessment; 951 (0.06%) and 775 (0.27%), respectively, also met the separately defined future persistent/recurrent-positive rule. More windows were eligible for persistent/recurrent prediction than for strict new onset, and eligibility for both decreased later in the ICU stay. These construction-induced quantities delimit the endpoint-specific cohorts; they are not false-positive rates or disease-incidence comparisons.

**Fig. 2.**
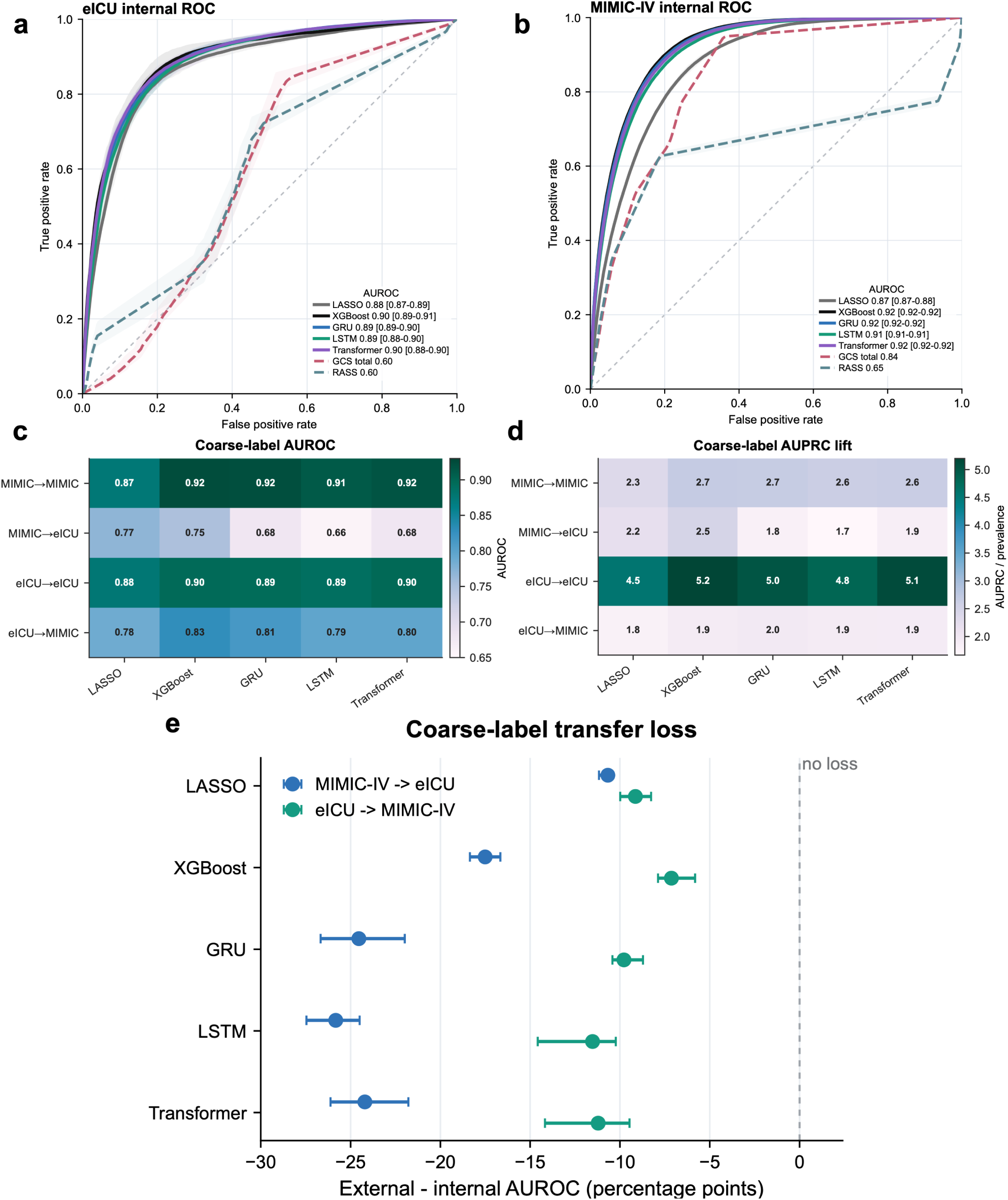
High internal coarse-label discrimination declined during source-only transfer. **a,b**, Internal receiver-operating-characteristic curves in eICU (**a**) and MIMIC-IV (**b**) for five model families; database-specific GCS- and RASS-based curves are shown as descriptive score references. Their score inputs and preprocessing differed by database, so they are not matched formal benchmarks. Solid curves show model ROC estimates and shaded bands show fold minima and maxima. **c**, Coarse-label AUROC for internal and source-only external validation. **d**, AUPRC lift, calculated as AUPRC divided by endpoint prevalence, for the same settings. **e**, Fold-paired external minus corresponding source-database internal AUROC in percentage points; values below zero indicate transfer loss. Points show the mean of five paired fold differences and horizontal intervals show the fold minimum–maximum range, not a confidence interval. Internal estimates are means over five folds and external estimates are five-fold ensembles.

**Fig. 3.**
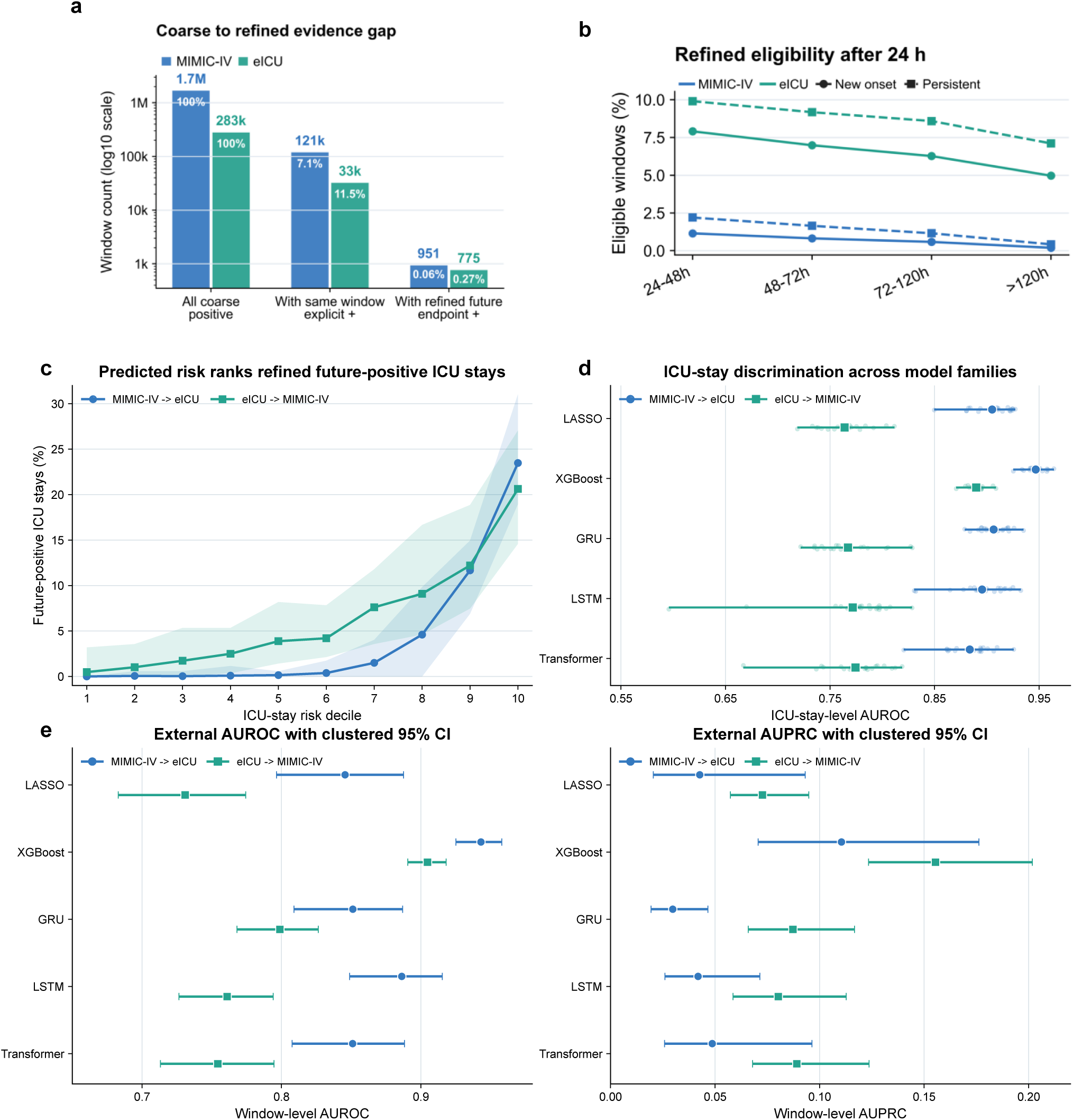
Endpoint construction altered eligibility and external model performance. **a**, Numbers of coarse-positive windows, those with a same-window explicit-positive assessment and those also meeting the separately defined future persistent/recurrent-positive rule. Percentages use all coarse-positive windows in the corresponding database as denominator. These construction-induced overlaps do not validate either label or estimate false-positive rates. **b**, Proportion of post-24 h windows eligible for strict new-onset (circles, solid lines) or persistent/recurrent (squares, dashed lines) operational targets across ICU-time strata. **c**, Future-positive ICU-stay rate by decile of maximum predicted GRU risk in source-only external validation. Lines show means across 20 model-training runs and shaded bands show the observed minimum–maximum range. **d**, ICU-stay-level AUROC based on each stay’s maximum predicted risk across five model families. Small points denote individual model-training runs, large symbols denote means and horizontal intervals denote observed minimum–maximum ranges across 20 runs. **e**, Window-level external AUROC (left) and AUPRC (right) for the full-clinical, both-history persistent/recurrent setting in one prespecified natural-person-grouped model and partition. Points show estimates and horizontal intervals show percentile 95% confidence intervals from 2,000 bootstrap replicates that resampled target ICU stays and retained all windows from each sampled stay. These intervals quantify target-cohort sampling uncertainty conditional on that model and partition; the ranges in **c,d** quantify training and split sensitivity and are not confidence intervals. Body-size-excluded and 20-run natural-person-grouped one-factor sensitivity summaries are reported in Supplementary Table S8; model-specific clustered estimates are tabulated in Supplementary Table S12. All targets are retrospective operational definitions rather than adjudicated diagnoses; their performance does not establish clinical superiority over alternative endpoint definitions.

Among ICU stays eligible for persistent/recurrent analysis, median age was 66 years in eICU and 65 years in MIMIC-IV, and 52.6% and 56.4%, respectively, were male (Supplementary Table S11). The first eligible anchor occurred at a median of 27 h in eICU and 38 h in MIMIC-IV, with a median of five and two eligible windows per stay. A positive assessment during the first 24 h was recorded in 14.7% and 32.2% of eligible stays, respectively. These descriptive differences further characterize the assessment-conditioned populations and were not used as tests of database equivalence.

### 2.3 Coarse-label signal was strongly learnable within each database

To test whether internal learnability implied cross-database transport, we evaluated LASSO, XGBoost, GRU, LSTM and Transformer models on the coarse-label benchmark. Internal AUROC ranged from 0.88 to 0.90 in eICU and from 0.87 to 0.92 in MIMIC-IV (Fig. 2a–c and Supplementary Table S10). The database-specific GCS- and RASS-based reference curves were lower, but their input and preprocessing implementations differed and were therefore descriptive rather than matched benchmarks. AUPRC lift over target prevalence ranged from 4.5 to 5.2 in eICU and from 2.3 to 2.7 in MIMIC-IV (Fig. 2d). No architecture was uniformly dominant across databases and discrimination metrics.

### 2.4 Source-only transfer exposed discrimination and temporal instability

Source-only transfer reduced discrimination for every model. Models trained in MIMIC-IV and evaluated in eICU achieved AUROCs of 0.66–0.77 and AUPRC lifts of 1.7–2.5, whereas eICU-to-MIMIC-IV transfer achieved AUROCs of 0.78–0.83 and AUPRC lifts of 1.8–2.0 (Fig. 2c,d). Every external-minus-corresponding-internal AUROC difference was negative, with larger reductions for MIMIC-IV-to-eICU transfer (Fig. 2e). High internal discrimination therefore did not identify a model family that was invariant to the target database.

Temporal sensitivity analyses crossed 5, 15, 30 and 60 min grids with 1–4 h horizons. Internal AUROC was comparatively stable at 15–60 min resolutions, whereas external results varied more across configurations and transfer directions; sequence models generally had the largest max–min ranges under transfer (Supplementary Figs. S1–S5). The transport deficit therefore persisted beyond the primary 60 min-grid, 4 h-horizon specification and was accompanied by greater temporal-configuration sensitivity externally than internally.

### 2.5 Persistent/recurrent risk remained externally rankable after integrity sensitivities

We next tested whether direct-assessment endpoint construction changed external learnability. For the operational persistent/recurrent target, mean future-positive ICU-stay rates in the highest GRU risk decile reached 23.5% for MIMIC-IV-to-eICU transfer and 20.6% for eICU-to-MIMIC-IV transfer (Fig. 3c). Across all five model families, mean ICU-stay-level AUROC was 0.88–0.95 and 0.76–0.89, respectively (Fig. 3d). By contrast, the strict new-onset sensitivity produced AUROCs of 0.50–0.58 across model families and 20 model-training runs per cell (Supplementary Fig. S6). This contrast establishes endpoint-specific learnability, not clinical superiority of one operational definition.

Two independent integrity sensitivities preserved this external pattern (Supplementary Table S8). When all ICU admissions from the same natural person were constrained to one partition, mean external AUROC ranged from 0.865 to 0.946 for MIMIC-IV-to-eICU transfer and from 0.770 to 0.904 for eICU-to-MIMIC-IV transfer. Relative to the primary ICU-stay-grouped analysis, cell-wise mean AUROC changed by −0.002 to 0.021 and AUPRC by −0.004 to 0.016. In a separate rerun excluding weight, height and body mass index, mean AUROC changed by −0.004 to 0.020 and AUPRC by −0.003 to 0.006. Each rerun changed one analysis factor while retaining the primary endpoint, full-clinical history specification, five model families, two transfer directions and 20 model-training runs per cell; they were not combined as a factorial analysis.

We separately quantified target-cohort sampling uncertainty for one prespecified person-grouped model and partition by resampling target ICU stays and retaining all windows from each sampled stay (Fig. 3e and Supplementary Table S12). Across the five models, window-level external AUROC ranged from 0.846 to 0.943 for MIMIC-IV-to-eICU transfer and from 0.731 to 0.905 for eICU-to-MIMIC-IV transfer; AUPRC ranged from 0.030 to 0.110 and from 0.073 to 0.155, respectively. For XGBoost, AUROC and AUPRC were 0.943 (95% CI, 0.925–0.958) and 0.110 (0.071–0.176) in the first direction and 0.905 (0.891–0.918) and 0.155 (0.123–0.202) in the second. These ICU-stay-clustered intervals describe sampling uncertainty conditional on that model and partition; the 20-run ranges in panels **c,d** continue to describe training and split sensitivity.

### 2.6 Assessment history dominated transported persistent/recurrent prediction

Input attribution identified longitudinal assessment and observation-process information as the dominant measured component of persistent/recurrent prediction (Fig. 4a). First-24 h and post-24 h delirium-history domains together accounted for 54.0% of total mean absolute SHAP attribution in MIMIC-IV-to-eICU transfer and 43.5% in eICU-to-MIMIC-IV transfer. ICU time contributed approximately 20% in each direction, and measurement timing or missingness contributed 12.0% and 13.9%, respectively. The remaining attribution was distributed across laboratory, hemodynamic, neurologic-assessment, demographic, medication, ventilation and care-setting domains.

**Fig. 4.**
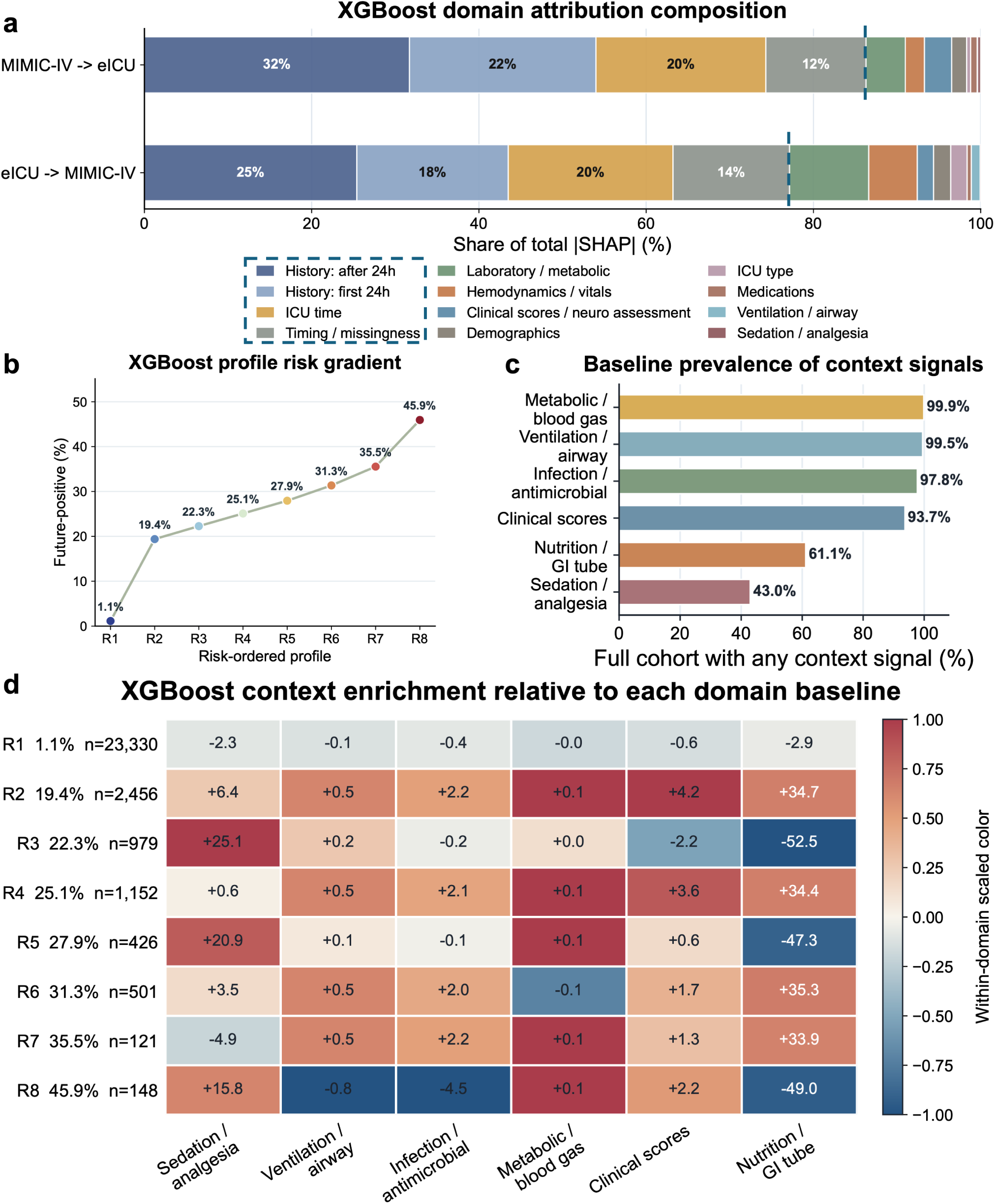
Transferred predictions emphasized assessment history and observation-process information. **a**, Share of total mean absolute SHAP value by input domain for full-clinical, both-history persistent/recurrent XGBoost models under bidirectional source-only external validation. Dashed boundaries separate the four dominant historical, ICU-time and observation-process domains from the remaining clinical domains. **b**, Future-positive rate across eight ICU-stay-level XGBoost probability-profile clusters, ordered post hoc from R1 to R8; cluster sizes are reported in panel **d**. **c**, Full-cohort ICU-stay prevalence of six curated clinical-context domains. **d**, Cluster-specific prevalence minus the corresponding full-cohort domain prevalence, in percentage points. Numeric entries are the unscaled differences; color is normalized separately within each domain to preserve contrast despite different baselines and therefore should not be compared as an absolute scale across columns. Probability-profile clusters were constructed from ICU-stay-level model outputs only. Context domains were joined afterward and were not used for clustering, training or SHAP computation.

History ablation directly tested whether this attribution translated into discrimination. In the full-clinical setting, the variant containing both first-24 h and post-24 h history achieved the highest mean AUROC for every model and validation direction (Fig. 5). Removing all assessment-history features reduced external AUROC by 0.24–0.32 for MIMIC-IV-to-eICU transfer and by 0.16–0.20 for eICU- to-MIMIC-IV transfer; retaining post-24 h history alone reduced AUROC by approximately 0.05–0.09 (Fig. 5a and Supplementary Fig. S16). Internally, removal of all history reduced AUROC by 0.09–0.15 in MIMIC-IV and by 0.16–0.25 in eICU (Fig. 5b and Supplementary Fig. S17). AUPRC lift generally followed the same ordering, although post-24 h-only history occasionally matched or modestly exceeded both-history performance for an individual model–metric combination.

**Fig. 5.**
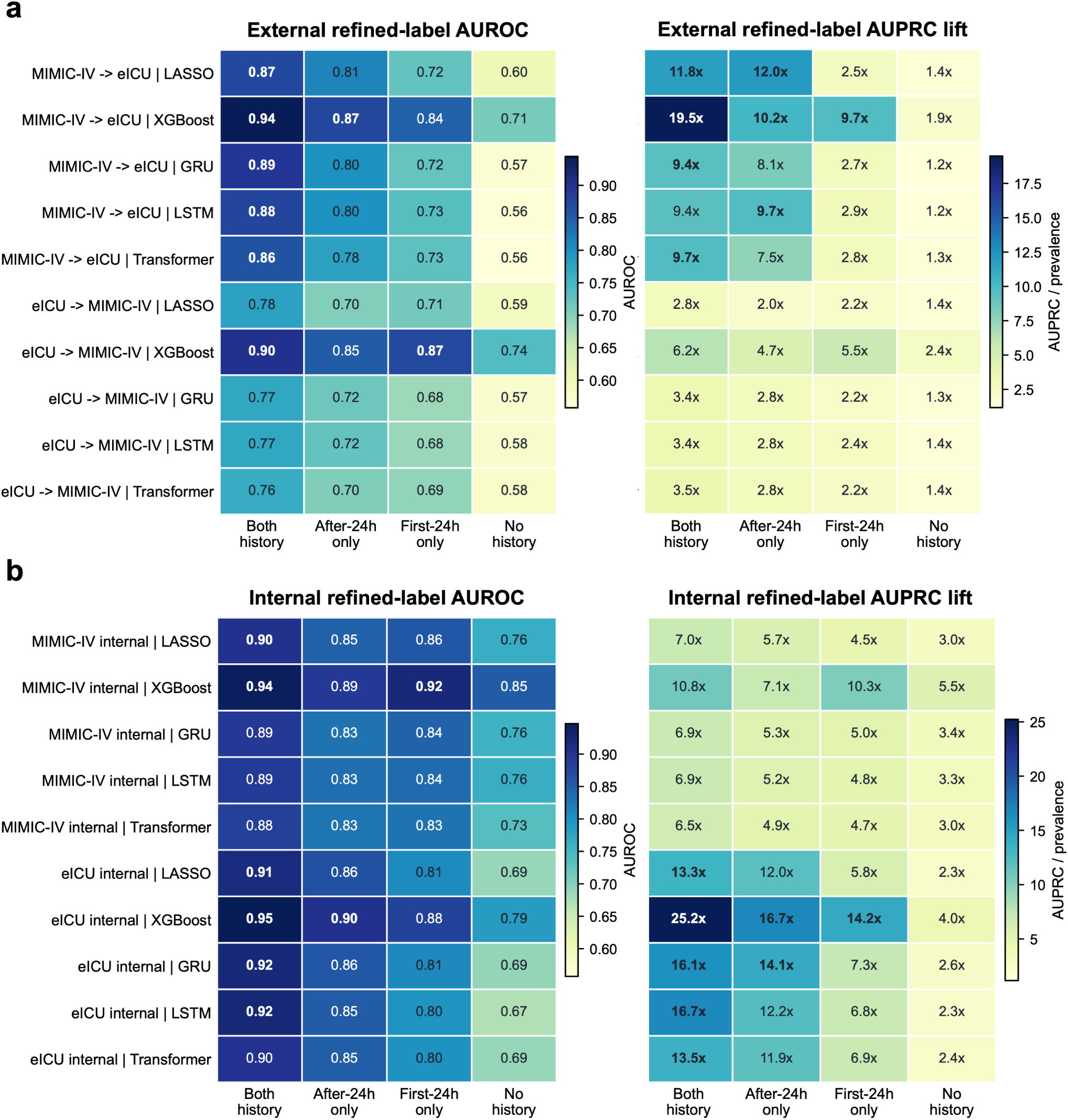
Assessment-history ablation markedly reduced persistent/recurrent discrimination. **a**, Mean external AUROC (left) and AUPRC lift (right) across 20 model-training runs for four feature-history variants: both first-24 h and post-24 h history, post-24 h history only, first-24 h history only and no assessment history. Rows identify transfer direction and model family. **b**, Corresponding internal-validation results. All panels use the full-clinical, 60 min-grid, 4 h-horizon setting. AUPRC lift is AUPRC divided by endpoint prevalence. The ablation measures dependence on available prior assessment information; a decrease after history removal is not, by itself, evidence of target leakage or clinical validity.

Assessment history did not exhaust the measurable signal. Refitting XGBoost without history redistributed mean absolute SHAP attribution toward neurologic or consciousness variables, measurement timing or missingness, laboratory measurements and other retained domains, while no-history models retained above-chance discrimination (Supplementary Fig. S18). Because these were separately fitted models, the redistribution does not estimate a causal substitution effect. It nevertheless shows that the operational target combined longitudinal label history with additional clinical and observation-process information.

### 2.7 Broader feature representations did not yield a uniform transport gain

Adding broader raw-source representations did not compensate consistently for history dependence. Expanded features increased GRU AUROC by 0.015 and 0.017 in the two transfer directions but changed Transformer AUROC by −0.022 and 0.002; expanded-feature AUPRC increased for LSTM in both directions but decreased for GRU and Transformer (Supplementary Figs. S12 and S13). Curated-allraw augmentation changed AUROC by at most 0.003 for XGBoost and the three sequence models but reduced LASSO AUROC by 0.089 and 0.062 (Supplementary Fig. S14); AUPRC changes ranged from −0.044 to 0.012 (Supplementary Fig. S15). More raw-source variables therefore produced model- and direction-specific changes rather than a general external-performance gain.

ICU-stay probability profiles further showed that transported risk was not reducible to a single post hoc clinical context. XGBoost profiles formed a gradient from 1.1% future-positive stays in R1 to 45.9% in R8 (Fig. 4b). Clinical-context variables were not used for clustering. Metabolic or blood-gas (99.9%), ventilation or airway (99.5%), infection or antimicrobial (97.8%) and clinical-score (93.7%) signals were nearly ubiquitous, whereas nutrition or gastrointestinal-tube and sedation or analgesia signals occurred in 61.1% and 43.0% (Fig. 4c). Context enrichment varied non-monotonically across profiles, with similar risk ordering and heterogeneous context for the other model families (Fig. 4d and Supplementary Figs. S7–S11). These profiles describe correlates of model outputs rather than causal phenotypes or care-action targets.

### 2.8 Supervised target adaptation improved transfer with sufficient target data

We finally tested whether target labels could improve transferred models and whether their scores retained operational meaning. Supervised target-label adaptation changed external window-level AUPRC in a data-size-dependent manner (Fig. 6a). Mean paired differences were heterogeneous at the 1% target-development setting, positive from 5% onward and largest when all target-development stays were used. The AUROC sensitivity followed the same data-size pattern: mean paired changes after full fine-tuning were negative with 1% of target-development stays, positive at 5–25% and largest at 100%, whereas head-only changes were smaller (Supplementary Fig. S19). Recalibration alone left ranking and AUROC unchanged whenever the fitted map was monotonic. Each adapted estimate was paired with its corresponding source-only model from the same initialization and transfer direction.

**Fig. 6.**
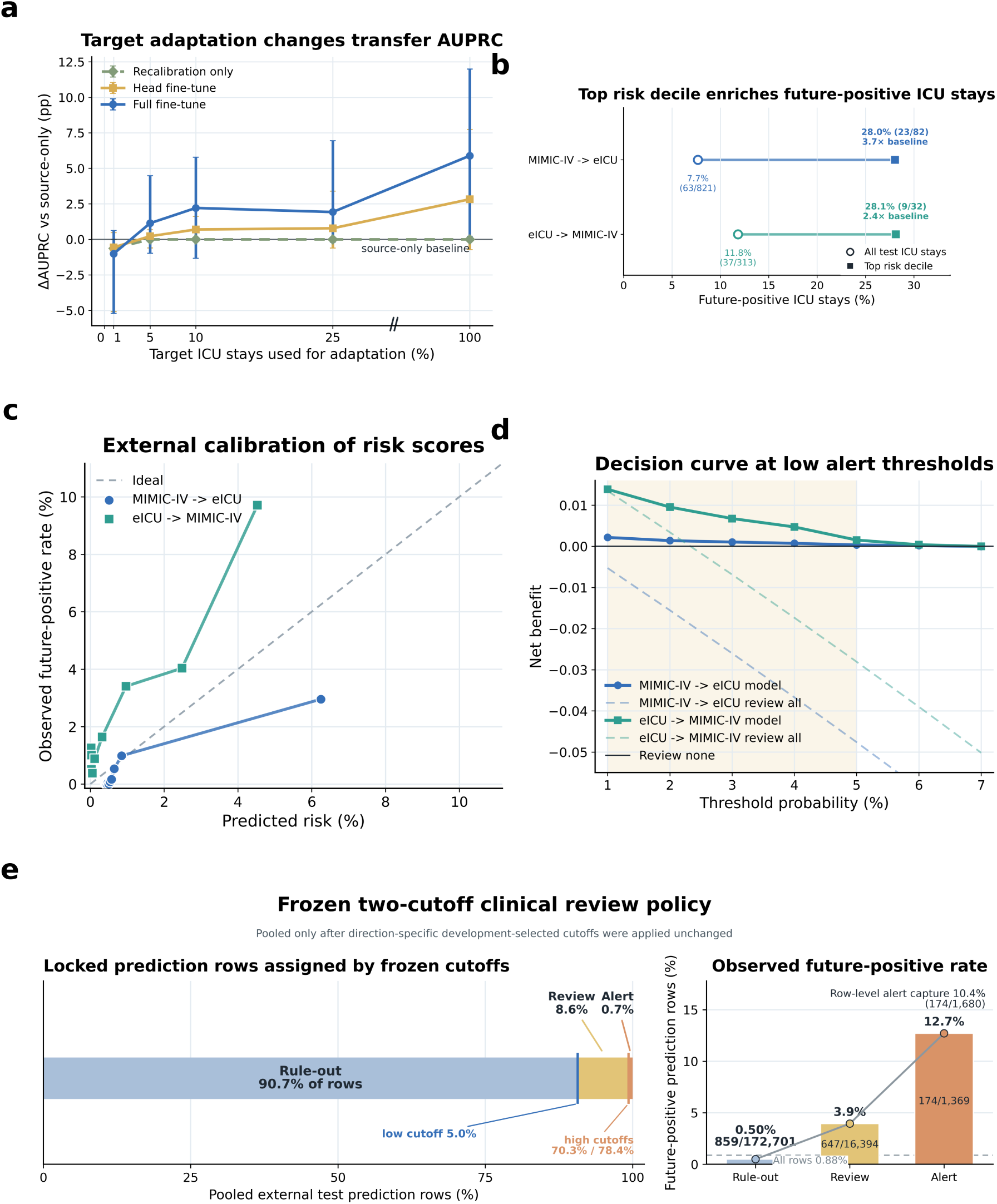
Supervised adaptation and frozen-threshold review analyses separate ranking from policy transport. **a**, External window-level AUPRC change relative to the paired source-only GRU as increasing fractions of labeled target ICU stays were used for recalibration or fine-tuning. Points show means and intervals show observed minimum–maximum ranges across eight direction–run results (four runs per direction), not confidence intervals. **b**, Future-positive proportion among ICU stays in the highest predicted-risk decile. Small points denote individual runs, large symbols denote means across 20 runs, intervals denote minimum–maximum ranges and open symbols denote overall rates. **c**, Direction-specific calibration bins for held-out external logistic-calibrated GRU scores; the diagonal denotes ideal calibration. Bins are descriptive for this sparse endpoint, and non-finite or non-positive calibration slopes were treated as instability flags. **d**, GRU window-level decision-curve net benefit at threshold probabilities of 1–7%. **e**, Frozen two-cutoff analysis for a prespecified source-only XGBoost model. Cutoffs were selected only from source-database internal development predictions and applied unchanged to external target-test prediction rows: 5.0% and 78.4% for MIMIC-IV-to-eICU, and 5.0% and 70.3% for eICU-to-MIMIC-IV. Assignments were pooled only after direction-specific threshold application; no common cutoff was selected from pooled test data. The band shows pooled prediction-row allocation, bars and points show zone-specific future-positive row rates, labels give positive/all rows, and the dashed line gives the overall row rate. The external-test data contained six prediction rows per unique source– target–ICU-stay–offset combination; deduplicated clinical-window, 8 h-shift, ICU-stay and refractory-period alert-episode workloads are reported in Supplementary Table S13. Direction-specific row-level estimates and 2,000-replicate fixed-threshold bootstrap intervals are in Supplementary Table S7. Target-label adaptation was supervised. Panels **c–e** are retrospective probability, review and alert-burden analyses; no prospective actionability, intervention effectiveness, workflow benefit or patient benefit was tested.

### 2.9 Risk ranking transported more consistently than probability and review policy

Even before local fine-tuning, transported scores concentrated future-positive ICU stays in the upper tail (Fig. 6b). Across 20 model-training runs per model and direction, the mean future-positive proportion in the top risk decile ranged from 23.1% to 29.6% for MIMIC-IV-to-eICU transfer and from 15.1% to 27.6% for eICU-to-MIMIC-IV transfer. Overall future-positive rates were approximately 4.2–4.3% and 6.3– 6.5%, yielding mean enrichment of 5.5–6.9-fold and 2.4–4.2-fold, respectively. Upper-tail prioritization was present across all five model families, although its magnitude was direction dependent.

Probability calibration did not share a common mapping across transfer directions (Fig. 6c). The upper eICU-to-MIMIC-IV calibration bin had a higher observed than mean predicted future-positive rate, whereas the upper MIMIC-IV-to-eICU bin showed the opposite pattern. Because future-positive labels were sparse, these bins were interpreted descriptively; non-finite or non-positive calibration slopes were treated as estimation-instability flags rather than evidence of perfect calibration. Over the displayed 1–7% threshold range, the logistic-calibrated GRU produced positive window-level decision-curve net benefit relative to review-none over most thresholds in both directions and exceeded review-all (Fig. 6d). These curves evaluated retrospective window-level review decisions rather than a deployed ICU-stay alert policy.

To evaluate a review rule without using external test labels for threshold selection, we selected direction-specific XGBoost cutoffs only from source-database internal development predictions and then applied them unchanged to external target-test prediction rows (Fig. 6e and Supplementary Table S7). For MIMIC-IV-to-eICU transfer, frozen cutoffs of 5.0% and 78.4% assigned 91.3% of 143,382 prediction rows to rule out, 8.4% to review and 0.3% to alert; observed future-positive rates were 0.18%, 2.84% and 13.78%, respectively. For eICU-to-MIMIC-IV transfer, cutoffs of 5.0% and 70.3% assigned 88.7% of 47,082 prediction rows to rule out, 9.2% to review and 2.0% to alert, with corresponding rates of 1.51%, 7.01% and 12.24%. After these direction-specific cutoffs had been applied, the compact display pooled the 190,464 prediction rows: 90.7%, 8.6% and 0.7% fell in the rule-out, review and alert zones, whose future-positive rates were 0.50%, 3.95% and 12.71%. This pooling was descriptive and did not re-estimate a common cutoff. Rule-out negative predictive values were 99.8% and 98.5% by direction, but the alert zones captured only 9.2% and 11.0% of future-positive prediction rows (10.4% pooled; 174 of 1,680).

The external-test prediction table contained six rows for every source–target–ICU-stay–offset combination. For workload estimation, we therefore collapsed 190,464 rows to 31,744 unique clinical windows and 4,548 ICU stays, assigning each deduplicated unit its highest risk zone (Supplementary Table S13). In MIMIC-IV-to-eICU and eICU-to-MIMIC-IV transfer, respectively, 1.1% and 7.3% of clinical windows and 4.9% and 12.2% of ICU stays entered the alert zone. Alerts captured 32.4% and 38.3% of future-positive clinical windows and 43.9% and 49.4% of future-positive ICU stays. Alert zones occurred in 1.9% and 8.2% of observed 8 h shifts. With an 8 h refractory period, 141 and 452 alert episodes remained, equivalent to 8.1 and 16.1 episodes per 100 ICU stays; 15.6% and 14.6% of episodes contained at least one future-positive alert window. Pooled totals were 827 alert clinical windows, 427 ICU stays with at least one alert and 593 alert episodes. These are retrospective workload and event-yield summaries, not independently adjudicated delirium episodes or evidence of prospective actionability, intervention effectiveness, workflow benefit or patient benefit.

## 3 Discussion

Cross-database transportability of ICU delirium prediction was layered rather than binary. Across five model families and two transfer directions, endpoint construction determined who and what could be predicted; source-only discrimination declined; persistent/recurrent ranking depended strongly on prior assessment history; and upper-tail risk concentration transported more consistently than probability calibration or review thresholds. Supervised local updating improved precision–recall performance once sufficient labelled target data were available. No architecture overcame the effects of the target data system, longitudinal documentation and the component of the prediction system being transported. The deployment question is not simply whether a model transports, but which of its endpoint, ranking, probability and policy layers remain valid at the target site.

Endpoint definition is the first layer because it fixes the clinical question before any model is trained. Carrying an observed state forward, forecasting a newly documented event and predicting persistence or recurrence impose different histories, eligible populations, event prevalences and opportunities for action. The low overlap between coarse-positive and future persistent/recurrent-positive windows therefore follows partly by construction; it neither estimates a false-positive rate nor establishes one operational target as closer to latent delirium. Its value is to show why performance from differently constructed delirium tasks cannot be compared as if it concerned one outcome. This distinction is especially important for a fluctuating syndrome observed intermittently through CAM/CAM-ICU or ICDSC assessments [1, 2, 5–7]. The near-chance strict new-onset sensitivity likewise defines a challenge for that endpoint and selected cohort, rather than a general claim that de novo delirium is unpredictable.

Within the operational persistent/recurrent task, assessment history was the largest measured component of transported ranking. Its use was temporally valid because all history preceded the anchor, and previous delirium is clinically prognostic for persistence or recurrence [27–29]. Yet prior assessments can jointly encode patient state, surveillance intensity, clinician suspicion, sedation and documentation practice. The large external AUROC reductions after history removal therefore indicate that these models partly monitored a documented longitudinal trajectory rather than estimating de novo biological onset from contemporaneous physiology alone. Above-chance no-history performance and redistributed attribution show that assessment history did not exhaust the signal, but they do not remove this dependence. Because eligibility also required direct assessment at the anchor and within the horizon, the estimand was conditional on the local observation process. The intended use is consequently repeated risk stratification among eligible, previously observed ICU stays, and the results should not be extrapolated to unassessed times or to population-wide delirium incidence.

The main transport pattern was not explained by repeated admissions from one person appearing in different partitions or by the three body-size variables. Natural-person-grouped and body-size-excluded reruns preserved above-chance discrimination across all five model families and both directions, with mean AUROC changes no larger than 0.021 and 0.020, respectively. These were separate one-factor sensitivities rather than a factorial reconstruction. More raw-source variables likewise did not produce a common transferable representation. Expanded and curated feature sets yielded model- and direction-specific changes, while SHAP attribution remained concentrated on assessment history, ICU time and measurement timing or missingness. Probability-profile clusters also spanned heterogeneous clinical contexts instead of resolving into discrete care-action phenotypes. These findings extend the progression from admission-time scores to nonlinear and recurrent delirium models by showing that greater representational complexity does not by itself solve cross-database shift [5, 6, 8, 10, 15]. External evaluation should therefore report the endpoint, predictor provenance and observation process alongside discrimination and calibration, rather than treating architecture rank as the principal evidence of portability [20, 22, 30].

Risk ranking, probability transport and policy transport also had different empirical behavior. Top-decile enrichment across all five model families showed that transferred scores could prioritize a smaller group with a higher future-positive rate, and supervised target adaptation improved AUPRC once additional labelled target stays were available. Development-selected cutoffs applied unchanged to external data preserved an ordered increase in future-positive rate from rule out to review to alert in both directions, but the selected cutoffs, zone allocation and event yield remained source–target dependent. At the prediction-row level, alerts captured only 9–11% of future-positive rows. After repeated rows were collapsed, 4.9–12.2% of ICU stays had at least one alert; an 8 h refractory rule yielded 8.1–16.1 alert episodes per 100 stays, of which 14.6–15.6% contained a future-positive alert window. Sparse outcomes also made calibration slope and intercept estimates unstable in some strata, so inference rested on ranking, descriptive calibration bins, decision curves, frozen-threshold operating characteristics and retrospective workload rather than a claim of perfect calibration or clinical benefit. Local updating, calibration and threshold selection should therefore be treated as separate implementation steps, followed by prospective evaluation of frozen policies, workflow, safety and downstream outcomes [21, 31, 32].

Taken together, these results recast ICU delirium-model transport as validation of a complete clinical prediction system rather than export of an architecture and an AUROC. The endpoint, eligible population, historical inputs, probability mapping and review policy each require target-site evaluation. In the present retrospective setting, the evidence supports cross-database risk stratification, supervised local updating and quantification of clinical-review and alert burden. It does not establish prospective actionability, intervention effectiveness, workflow benefit or improved patient outcomes. A prospective study can now test a fully specified endpoint, model, calibration map and review workflow unchanged across additional hospitals. This layered standard makes clear both what has transported and what must still be established before an ICU delirium prediction system can guide care.

## 4 Methods

### 4.1 Study design and prediction framework

We performed a retrospective cross-database observational study using the de-identified critical-care EHR databases eICU version 2.0 and MIMIC-IV version 3.1 [25, 26, 33, 34]. The study did not prospectively assign participants to an intervention and was not a clinical trial. The eICU release contains admissions recorded in 2014–2015 across 335 units at 208 hospitals in the United States, whereas MIMIC-IV contains admissions recorded in 2008–2022 at Beth Israel Deaconess Medical Center in Boston, Massachusetts. These sources provided a deliberate contrast between a multicentre tele-ICU-derived database and a single-centre academic health-system database. ICU stays were the unit of longitudinal analysis, and database-specific event times were converted to ICU-relative offsets before feature and label harmonization. The study evaluated how label construction affected the prediction task, how models transported bidirectionally between databases, which longitudinal information contributed to transported predictions, and how local adaptation and threshold-based review changed retrospective deployment-facing performance.

For ICU stay *i*, grid time *t_ij_* = *j*Δ paired all feature information available by the anchor with an outcome observed during the subsequent horizon:

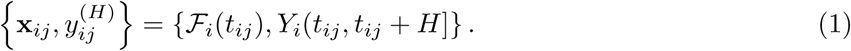

The primary refined setting used Δ = 60 min, *H* = 4 h and anchors at or after 24 h. Future assessments defined outcomes only and were never included in anchor-time predictors. The primary endpoint was persistent or recurrent delirium within the next 4 h; strict new onset was evaluated as a harder endpoint sensitivity.

Internal validation trained and evaluated within one database. Source-only external validation trained in one database and evaluated the held-out test partition of the other without target-database retraining. Bidirectional transfer was evaluated from eICU to MIMIC-IV and from MIMIC-IV to eICU. The same primary refined specification was used for Figs. 3–5; temporal, endpoint, feature and history variants were analyzed separately. Cohort construction, grid eligibility and database-specific source counts are reported in Supplementary Methods and Supplementary Tables 1 and 2.

No formal sample-size calculation was performed because the study analyzed fixed retrospective database releases. We included all ICU stays satisfying the prespecified temporal-grid and endpoint-specific eligibility rules; achieved ICU-stay, window and future-positive-event counts are reported in Supplementary Table S1 and Supplementary Fig. S23.

### 4.2 Delirium observations and endpoint construction

Each grid cell was classified as explicit positive, explicit negative, conflict, unable to assess or no assessment. Explicit positive and conflict states supplied positive point evidence, explicit negative supplied negative point evidence, and unable-to-assess or absent records were not converted to negative labels. eICU assessments were harmonized from nursing-charted delirium fields, including CAM-ICU- and ICDSC-related records; MIMIC-IV assessments were harmonized from corresponding charted items. Valid ICDSC values of 4 or greater were positive and lower values were negative. Deep sedation, defined as RASS ≤ −4 when available without direct positive or negative delirium evidence, was unable to assess [3, 4, 35]. These assessments had been documented during routine care before retrospective model development and were not generated by the study models. Individual assessor identities, qualifications and demographic characteristics were not retained in the de-identified releases.

The coarse benchmark carried the latest direct point label forward within each stay. Let *A_i_*(*t*) indicate direct assessment, *Z_i_*(*t*) ∈ {0, 1} its point label, and *s_i_*(*t*) = max{*s ≤ t*: *A_i_*(*s*) = 1}. The carried state and horizon target were

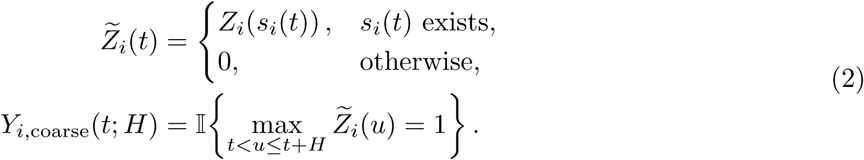

A positive state could therefore continue until a later negative assessment or ICU discharge. We treated this as a deterministic weak-label benchmark, not as an adjudicated future event.

Refined endpoints required direct assessment evidence. Strict new-onset anchors had a current explicit-negative state, no earlier positive point evidence and at least one direct assessment during the horizon; the target was positive only if that future assessment was positive. For persistent/recurrent delirium, positive points were grouped into episodes. A point was classified as persistent/recurrent, *P_i_*(*t*) = 1, when its observed positive episode had spanned at least 48 h or when it belonged to a second or later episode separated by at least 48 h. Eligible anchors required current and future direct assessment and no persistent/recurrent event at or before the anchor:

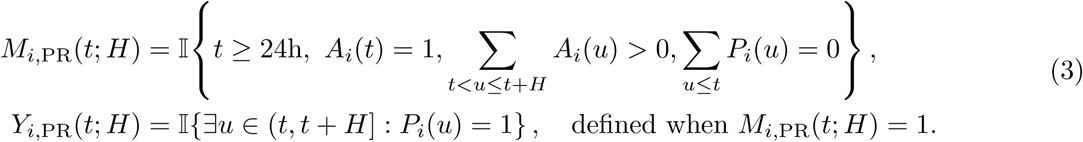

The resulting estimand was assessment conditioned: models estimated Pr{*Y_i,_*_PR_(*t*; *H*) = 1 | *M_i,_*_PR_(*t*; *H*) = 1*, F_i_*(*t*)}, rather than population-wide delirium risk at every ICU time. It therefore represented repeated monitoring among ICU stays with documented direct assessment at the anchor and during the future horizon. The exact state mapping, episode algorithm, strict new-onset mask and label denominators are specified in Supplementary Methods.

### 4.3 Features, model families and validation

The prespecified harmonized representation included demographics and ICU context, vital and respiratory measurements, laboratory measurements, neurologic or bedside scores, medication and vasoactive exposures, respiratory support, and observation-process variables. Numeric variables used the latest value in a prespecified lookback, an observation indicator and time since last observation; interval variables indicated any overlap with the lookback. Vital, respiratory and neurologic variables used 6 h lookbacks, laboratory variables 24 h, and medications, support and process events 6 h unless otherwise specified. Treatment and support variables were used only as time-indexed predictors in this observational prediction study; no treatment effects were estimated. The full-clinical representation was used for primary refined analyses. A no-clinical-score sensitivity representation removed RASS-, GCS-, pain-, APACHE-, OASIS- and related score fields. A separate body-size-excluded sensitivity removed weight, height and body mass index, reducing the full-clinical representation from 137 to 134 features while retaining the primary endpoint and history specification. Persistent/recurrent analyses compared no history, first-24 h history only, post-24 h history only and both-history variants.

We evaluated LASSO logistic regression, XGBoost, GRU, LSTM and Transformer models [36–40]. In the primary analysis, ICU stays were partitioned within each database into 70% training, 15% validation and 15% test sets, with all windows from a stay kept together. A natural-person-grouped sensitivity instead constrained all eICU admissions sharing an upstream uniquepid and all MIMIC-IV admissions sharing a subject id to the same partition. Numeric predictors were median-imputed and standardized using training data. LASSO provided a sparse linear benchmark and XGBoost represented nonlinear tabular effects. GRU, LSTM and Transformer models received causal, chronologically ordered anchor sequences; future rows and padded positions were excluded from prediction and loss. Neural models used class-weighted binary cross-entropy, validation-based early stopping and checkpoint selection. Model dimensions, optimization settings and the descriptive GCS/RASS reference implementations are reported in Supplementary Methods and Supplementary Table 4.

Coarse-label analyses used fivefold ICU-stay-level cross-validation and the corresponding source ensembles for external evaluation. Primary refined analyses used 20 model-training runs per model and setting. A model-training run denoted one complete fit with a distinct initialization; cross-validation folds, bootstrap resamples and clustering initializations were separate procedures.

### 4.4 Attribution, profiles and sensitivity analyses

For external XGBoost models, TreeSHAP values were computed on held-out target rows and mean absolute values were summed within prespecified feature domains. Shares were normalized within each transfer direction and were interpreted as fitted-model dependence rather than causal effects. Separately, each ICU stay was summarized by its mean and maximum predicted probabilities across 20 runs. Standardized probability summaries were clustered by K-means into eight groups without using outcomes or clinical-context variables; R1–R8 labels were assigned post hoc by increasing future-positive rate. Clinical-context prevalence and enrichment were calculated only after clustering. t-SNE and UMAP were descriptive projections and did not determine cluster assignments.

Sensitivity analyses crossed four grid resolutions (5, 15, 30 and 60 min) with four horizons (1– 4 h), compared strict new-onset with persistent/recurrent endpoints, expanded the clean feature set with process and treatment proxies, evaluated manually curated raw-source candidates after excluding assessment-adjacent variables, ablated the two assessment-history blocks, grouped admissions by natural person and excluded the three body-size features. The person-grouped and body-size-excluded analyses were separate one-factor reruns of the five-model, two-direction, 20-run primary refined analysis. Feature selection or curation used no validation or test labels. Detailed attribution, clustering, feature-screening and sensitivity procedures are provided in Supplementary Methods.

### 4.5 Calibration, target adaptation and retrospective review policies

Discrimination was summarized by AUROC and AUPRC; AUPRC was also divided by evaluation prevalence to report lift over random ranking. Logistic calibration was fit on validation predictions and applied unchanged to held-out test predictions [31, 41]. Calibration intercept and slope, Brier score, log loss and calibration bins assessed probability accuracy. Because future-positive labels were sparse, calibration curves were treated as descriptive binned summaries; non-finite or non-positive slope estimates were flagged as calibration-model failure or instability and were not interpreted as perfect calibration. Prespecified single thresholds maximized validation recall subject to a source-validation false-positive rate no greater than 5%, with 10% as an alert-burden sensitivity. Decision-curve analysis compared model-guided review with review-none and review-all policies [21].

Target-label adaptation was supervised. Source models were updated with labeled target-development ICU stays at fractions 0, 1, 5, 10, 25 and 100%; zero denoted source-only transfer. Calibration-only adaptation kept the model fixed, head-only fine-tuning updated the classifier, and full fine-tuning updated all trainable parameters. Adaptation subsets were sampled by ICU stay, and each adapted estimate was paired with the source-only estimate from the same run and direction. Figure 6a,c,d used the persistent/recurrent, 60 min/4 h, no-clinical-score, both-history GRU setting; panel b compared all five model families, and panel e used the prespecified source-only XGBoost model described below. Additional settings are reported in Supplementary Methods.

For the final two-cutoff analysis, each external-test XGBoost prediction row *r*, associated with ICU stay *i* and offset *t_ij_*, was assigned to one of three zones:

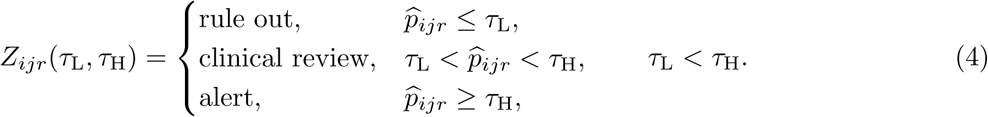

For each transfer direction, cutoffs were selected only from the source database’s internal development rows, targeting rule-out NPV 0.95 and alert specificity 0.95 with a low-cutoff ceiling and high-cutoff floor of 0.05. The selected pair was then frozen and applied unchanged to held-out external target-test prediction rows. The prespecified XGBoost model yielded (*τ*_L_*, τ*_H_) = (0.050, 0.784) for MIMIC-IV-to-eICU transfer and (0.050, 0.703) for eICU-to-MIMIC-IV transfer. Figure 6e pooled row-level zone counts only after the corresponding direction-specific pair had been applied; no common high cutoff was estimated from pooled test labels.

The external-test prediction table contained six rows per source–target–ICU-stay–offset combination. For the workload analysis, rows sharing this key were collapsed to one clinical window using the maximum risk-zone rank, with rule out < review < alert; the window outcome was the maximum existing row label. Clinical windows were then collapsed by maximum zone and label within each 8 h ICU-stay shift and within each ICU stay. An alert episode began at the first alert clinical window, and subsequent alert windows before a fixed refractory interval had elapsed from episode onset were grouped into that episode. Eight hours was the primary refractory period, with 4 h and 12 h sensitivities. No model was retrained and no threshold was reselected. These analyses quantified retrospective workload and event yield; unit outcomes were not independently adjudicated delirium episodes, and the analysis did not test prospective actionability, an intervention, workflow benefit or patient benefit. Direction-specific row-level estimates and fixed-threshold bootstrap intervals are reported in Supplementary Table S7; deduplicated workload is reported in Supplementary Table S13. Same-test-selected ICU-stay results are reported separately as exploratory Supplementary sensitivity analyses.

### 4.6 Statistical analysis

Descriptive quantities were calculated at the window or ICU-stay level stated in each figure. Assessment counts per ICU-day were compared between databases with a two-sided, tie- and continuity-corrected asymptotic Mann–Whitney *U* test. Coarse internal performance was summarized across five folds, and transport differences paired each external fold ensemble with its source-internal counterpart. Refined figures displayed individual model-training runs with their mean and observed minimum–maximum range unless stated otherwise; these ranges were not confidence intervals or independent biological replicates. Figure 3e instead reports percentile 95% intervals from ICU-stay-clustered bootstrap resampling of one prespecified person-grouped model and partition. We did not use pairwise significance tests to rank model families.

Except where stated otherwise, confidence intervals were obtained by bootstrap resampling ICU stays to preserve within-stay dependence. The frozen prediction-row two-cutoff operating summaries used percentile 95% intervals from 2,000 bootstrap replicates with thresholds held fixed. The deduplicated workload analysis was descriptive and did not use confidence intervals. The exploratory same-test-selected ICU-stay analysis used 1,000 ICU-stay bootstrap resamples and is reported only in the Supplementary Information. All reported tests were two sided, and no multiplicity adjustment was applied to descriptive sensitivity analyses. Formulae for calibration, net benefit, attribution shares, profile enrichment and fold-paired transport differences are given in Supplementary Methods.

To distinguish target-cohort sampling uncertainty from model-training and split variation, a prespecified sensitivity selected one natural-person-grouped model and partition before generating 2,000 percentile bootstrap replicates separately for each model and transfer direction. Target ICU stays were sampled with replacement and all prediction windows from each sampled stay were retained. These intervals were conditional on the selected model and partition and therefore complemented, rather than replaced, the observed ranges across 20 model-training runs.

## Ethics and consent

Both source resources contain retrospective records that were de-identified before distribution and were accessed through PhysioNet credentialing under the applicable data-use agreements. The collection of patient information and creation of MIMIC-IV as a research resource were reviewed by the Institutional Review Board at Beth Israel Deaconess Medical Center, which granted a waiver of informed consent and approved the data-sharing initiative [26, 34]. eICU-CRD was de-identified to the safe-harbor standard of the US Health Insurance Portability and Accountability Act; direct identifiers were removed, record identifiers were randomized and no linkage key to the original identifiable records was retained [25, 33]. The present analyses used only the distributed de-identified records and involved no patient contact.

## Supplementary information

Supplementary Information accompanies this manuscript.

## Data Availability

This study used the eICU Collaborative Research Database version 2.0 and MIMIC-IV version 3.1. Both datasets are available through PhysioNet to credentialed users who complete the required human-research training and sign the applicable data-use agreement (eICU: https://doi.org/10.13026/C2WM1R; MIMIC-IV: https://doi.org/10.13026/kpb9-mt58).

