## Supplemental information for "Cross-database validation reveals distinct layers of transportability in ICU delirium prediction"

### S1 Supplementary Methods

The main Methods retain the study estimands, primary endpoint, model families and evaluation framework. This section provides the full operational definitions, implementation details and mathematical summaries used to reproduce the analyses.

#### S1.1 Cohort construction and temporal-grid eligibility

ICU stays were indexed at the stay level, and event times were converted to ICU-relative offsets. Complete stay grids extended from ICU admission to ICU discharge or to the last assessment-derived follow-up when discharge time was unavailable. For stay $i$, grid width $\Delta$, anchor $t_{ij} = j\Delta$ and horizon $H$, each analysis row was

$$\left\{\mathbf{x}_{ij}, y_{ij}^{(H)}, M_{ij}^{(H)}\right\} = \{\mathcal{F}_i(t_{ij}), Y_i(t_{ij}, t_{ij} + H], M_i(t_{ij}; H)\}, \tag{S1}$$

where $M_{ij}^{(H)}$ was the endpoint-specific eligibility mask. Model fitting and evaluation used only rows with $M_{ij}^{(H)} = 1$. The primary refined analysis used a 60 min grid, excluded the first 24 h from anchor eligibility and evaluated a 4 h future horizon. All rows from one ICU stay remained in one training, validation or test partition. In the natural-person-grouped sensitivity, every ICU stay sharing the same upstream person identifier was additionally constrained to one partition.

The analysis grid contained 28,321 eICU stays and 2,496,432 hourly windows and 49,828 MIMIC-IV stays and 5,881,343 hourly windows. Of these, 24,060 eICU stays contributed 1,845,924 post-24 h windows and 47,181 MIMIC-IV stays contributed 4,694,559 post-24 h windows. Direct explicit assessments were present in 380,780 windows from 27,216 eICU stays and 457,471 windows from all 49,828 MIMIC-IV stays; after 24 h, the corresponding counts were 261,345 windows from 20,387 eICU stays and 333,261 windows from 42,425 MIMIC-IV stays. The strict new-onset endpoint was defined for 117,519 windows from 9,131 eICU stays, including 1,067 positive windows, and 26,405 windows from 9,823 MIMIC-IV stays, including 854 positive windows. The persistent/recurrent endpoint was defined for 157,056 windows from 11,572 eICU stays, including 782 positive windows, and 52,530 windows from 18,736 MIMIC-IV stays, including 1,224 positive windows. These model-grid and endpoint-eligibility counts are reported at ICU-stay and window levels. The primary specification used a 60 min resolution, 4 h horizon, 24 h minimum anchor offset and a minimum of three distinct grid cells containing explicit positive or negative assessment evidence per ICU stay. The label-grid builder applied no additional age or minimum ICU-duration filter beyond the source tables. Grids began at ICU admission and ended at recorded ICU discharge; when discharge time was unavailable, the last mapped assessment plus the 4 h horizon defined the fallback end time.

#### S1.2 Delirium assessment source mapping

Direct delirium evidence was extracted from database-specific structured sources and harmonized to explicit positive, explicit negative, conflict, unable to assess or no assessment. In eICU, candidate rows were identified when nursing-chart labels or names contained delirium, confusion, CAM-ICU or ICDSC terms. Normalized positive, present, yes, 1 or true values were mapped to explicit positive; negative, absent, no, 0 or false values were mapped to explicit negative; and unable, UTA, not assessed, cannot assess, not applicable or unknown values were mapped to unable to assess. Valid numeric ICDSC values from 0 to 8 were positive at 4 or greater and negative below 4. The source mapping contained 44,982 explicit-positive, 459,043 explicit-negative, 30,806 unable-to-assess and 293,266 unmapped or header-like eICU candidate rows before hourly collapse.

In MIMIC-IV, chart events with item identifiers 228332 and 229326 were aligned to ICU-relative time and mapped with the same normalized binary or textual rules. Item 228332 contributed 118,632 explicit-positive, 358,477 explicit-negative and 129,796 unable-to-assess rows; item 229326 contributed 134,249 explicit-positive and 207,012 explicit-negative rows before hourly collapse. RASS item 228096 was used only to identify deep sedation. A grid cell with RASS less than or equal to $-4$ and no direct positive or negative delirium evidence was unable to assess rather than negative.

Multiple raw rows within a grid cell were collapsed by evidence priority. Simultaneous positive and negative evidence produced a conflict state. Conflict cells were retained as flagged direct evidence and counted as positive point evidence for endpoint construction. Unable-to-assess and no-assessment cells were not converted to negative labels. Raw-row mapping counts describe provenance and are not independent assessments or ICU stays.

#### S1.3 Formal endpoint masks and label-family denominators

Let $A_i(t) = 1$ denote a direct assessment at grid time $t$, and let $Z_i(t) \in \{0, 1\}$ be its point label, with explicit positive or conflict mapped to 1 and explicit negative to 0. The coarse weak label used the latest direct point label within each stay. If $s_i(t) = \max\{s \leq t : A_i(s) = 1\}$, then

$$\begin{aligned} \widetilde{Z}_i(t) &= \begin{cases} Z_i(s_i(t)), & s_i(t) \text{ exists}, \\ 0, & \text{otherwise}, \end{cases} \\ Y_{i,\text{coarse}}(t; H) &= \mathbb{I}\left\{ \max_{t<u\leq t+H} \widetilde{Z}_i(u) = 1 \right\}. \end{aligned} \tag{S2}$$

This construction allowed a direct positive state to persist until the next observed negative assessment or ICU discharge.

Strict new-onset anchors required a current direct-negative state, no prior positive point evidence, at least one future direct assessment and $t \geq 24$ h. The eligibility mask and target were

$$\begin{aligned} M_{i,\text{NO}}(t; H) &= \mathbb{I}\left\{ \begin{array}{c} t \geq 24\text{h}, \quad A_i(t) = 1, \quad Z_i(t) = 0, \\ \sum_{u<t} \mathbb{I}\{A_i(u) = 1, Z_i(u) = 1\} = 0, \quad \sum_{t<u\leq t+H} A_i(u) > 0 \end{array} \right\}, \\ Y_{i,\text{NO}}(t; H) &= \mathbb{I}\{\exists u \in (t, t+H] : A_i(u) = 1, Z_i(u) = 1\}, \end{aligned} \tag{S3}$$

with $Y_{i,\text{NO}}$ defined only when $M_{i,\text{NO}} = 1$. This kept previously positive stays from re-entering the strict new-onset risk set and distinguished an observed-negative horizon from a horizon with no assessment.

For persistent/recurrent delirium, let $p_{i1} < \cdots < p_{iK_i}$ be positive grid times and $e_{ik}$ their episode indices. A gap of at least $G = 48$ h initiated a new episode. If $a_{ir}$ was the first positive time in episode $r$, a positive point became persistent/recurrent when its within-episode observed-positive span reached $D = 48$ h or it belonged to episode 2 or later:

$$P_i(t) = \mathbb{I}\{\exists k : p_{ik} = t,\ [p_{ik} - a_{i,e_{ik}} \geq D \text{ or } e_{ik} \geq 2]\}, \qquad D = G = 48\text{h}. \tag{S4}$$

The primary mask and target were

$$\begin{aligned} M_{i,\text{PR}}(t; H) &= \mathbb{I}\left\{ t \geq 24\text{h},\ A_i(t) = 1, \sum_{t<u\leq t+H} A_i(u) > 0, \sum_{u\leq t} P_i(u) = 0 \right\}, \\ Y_{i,\text{PR}}(t; H) &= \mathbb{I}\{\exists u \in (t, t+H] : P_i(u) = 1\}, \quad \text{defined only when } M_{i,\text{PR}}(t; H) = 1. \end{aligned} \tag{S5}$$

These rules define operational EHR endpoints rather than expert-adjudicated clinical episodes.

The coarse-versus-refined overlap analysis used the common 60 min/4 h setting. Among coarse-positive windows, we calculated the proportions with any same-window direct assessment, a same-window explicit-positive label and a future persistent/recurrent refined-positive target. ICU-stay overlap compared the presence of any coarse-positive state with any refined future-positive target. These quantities were label-construction diagnostics and were not used for model training.

#### S1.4 Feature harmonization, lookbacks and history variants

Harmonized features comprised demographics and unit context, vital and respiratory measurements, laboratory measurements, neurologic or bedside scores, medication and vasoactive exposures, respiratory support and process-sensitive variables. For numeric feature $k$ with lookback $L_k$, let $s^*_{ik}(t) = \max\{s \in (t - L_k, t] : k \text{ observed at } s\}$. The anchor representation retained

$$x_{ik}(t) = \begin{cases} v_{ik}(s^*_{ik}(t)), & s^*_{ik}(t) \text{ exists}, \\ \text{NA}, & \text{otherwise}, \end{cases} \quad o_{ik}(t) = \mathbb{I}\{s^*_{ik}(t) \text{ exists}\}, \quad r_{ik}(t) = t - s^*_{ik}(t), \tag{S6}$$

where $x$, $o$ and $r$ were the latest value, observation indicator and recency. Interval features used any overlap with the lookback. Vital, respiratory and neurologic variables used 6 h lookbacks, laboratory variables used 24 h, and medication, vasoactive, respiratory-support and process-event indicators used 6 h unless specified otherwise.

The primary refined analyses used the full-clinical feature set. The no-clinical-score sensitivity removed RASS-, GCS-, pain-score-, APACHE-, OASIS- and related score fields. Primary persistent/recurrent `both_hist` tables contained 137 full-clinical and 124 no-clinical-score feature columns. The body-size-excluded sensitivity removed only `weight`, `height` and `bmi`, leaving 134 full-clinical features; partitioning, endpoint construction, history features and model settings otherwise followed the primary analysis. Four history variants were evaluated: `no_hist`, `only_1st_hist`, `only_after_hist` and `both_hist`. History blocks contained counts, indicators and recency summaries of direct, positive, negative and unable-to-assess states. When no prior post-24 h direct assessment existed, recency was set to 168 h and paired with an indicator denoting that absence. Supplementary Data 1 provides per-column source, construction, lookback, missingness handling, feature-set membership and database-specific observed rate.

#### S1.5 Feature-expansion and raw-source sensitivity analyses

Expanded features added selected medication, culture, imaging, procedure and measurement-process indicators to the prespecified clean representation. Separately, candidate-feature associations were estimated in training data only within each database; stable top-ranked candidates defined association-selected subsets evaluated in five model-training runs. No validation or test labels were used for feature selection.

A raw-source review screened laboratory, vital-sign, charted-assessment, medication, infusion, treatment, procedure, microbiology, input and prescription tables where available. Manual curation excluded label-proximal, mental-status, CAM-ICU, GCS-verbal and assessment-adjacent candidates. Remaining candidates were grouped into core physiology, respiratory or ventilation detail, treatment or medication proxies, hemodynamic or invasive monitoring, infection or workup processes, nutrition or tube-care signals and clinical-action signals. Expanded and curated-allraw analyses were supplementary sensitivities and were not used to select the primary model representation.

#### S1.6 Model training, partitions and hyperparameters

ICU stays were assigned within each database to 70% training, 15% validation and 15% test partitions. The primary analysis grouped rows by ICU stay. For the natural-person-grouped sensitivity, eICU `patientunitstayid` values were mapped to `uniquepid` from the upstream patient table and MIMIC-IV stay identifiers were mapped to `subject_id` from the upstream ICU-stay table; ICU-stay identifiers were not treated as natural-person identifiers. All admissions sharing a mapped person identifier were assigned together, and split verification found no person crossing training, validation and test partitions. Tabular predictors were median-imputed and standardized using source-training statistics. LASSO used class-balanced weighting, an $L_1$-penalized logistic objective, the `saga` solver and at most 3,000 iterations. XGBoost used binary logistic loss, histogram tree construction, 500 estimators, maximum depth 3, learning rate 0.03, subsampling 0.85, column subsampling 0.85, minimum child weight 5, $L_2$ regularization 2.0 and positive-class weighting based on the source-training class ratio.

GRU, LSTM and Transformer models received chronologically ordered anchor sequences with the imputed and standardized predictors and previous-anchor changes. Primary refined runs used hidden or embedding dimension 96, one recurrent or Transformer layer, dropout 0.25, batch size 16, maximum sequence length 512, AdamW learning rate $10^{-3}$, weight decay $10^{-4}$ and weighted binary cross-entropy with positive-class weight capped at 100. The Transformer used four attention heads, feed-forward dimension 256, sinusoidal positional encoding and a causal attention mask. Source training used at most 30 epochs, at least 6 epochs before early stopping and patience 5; target fine-tuning used at most 20 epochs. Gradient norm was clipped at 5.0. Sequences exceeding the maximum length were split chronologically, and padded positions were excluded from loss and output.

A cross-validation fold referred to one held-out partition. A model-training run referred to one complete fit with a distinct initialization, and a direction–run result to evaluation of that fit in one transfer direction.

Bootstrap resampling and clustering initializations were separate procedures. Coarse analyses used fivefold cross-validation, whereas primary refined model-family and history-variant analyses used 20 model-training runs per analysis cell.

The GCS- and RASS-based curves in main Fig. 2a,b were database-specific descriptive score references rather than harmonized benchmarks. The eICU implementation used the current score plus a within-stay first difference, complete-analysis-table mean imputation before fold separation, training-fold standardization and class-balanced logistic regression. The MIMIC-IV implementation used the current score alone with training-fold median imputation, training-fold standardization and class-balanced logistic regression. Both used fivefold ICU-stay-stratified cross-validation. Their inputs and preprocessing boundaries differed, so no inferential or superiority claim relied on these references.

#### S1.7 Input attribution and probability-profile construction

Direction-specific XGBoost TreeSHAP values were calculated for held-out target rows in the full-clinical, `both_hist` setting. If $\mathcal{K}_d$ denotes features in domain $d$, $\mathcal{K}$ all features and $\overline{|\phi_k|}$ the target-set mean absolute SHAP value for feature $k$, the displayed domain share was

$$S_d = 100\,\frac{\sum_{k\in\mathcal{K}_d}\overline{|\phi_k|}}{\sum_{k\in\mathcal{K}}\overline{|\phi_k|}}. \tag{S7}$$

Shares were normalized separately by direction and quantify fitted-model dependence, not sign or causal effect.

For each run and transfer direction, window probabilities were summarized for each ICU stay by their maximum and mean, then averaged across 20 model-training runs. Available direction-specific components were median-imputed, standardized and clustered by K-means with eight clusters and 20 initializations. K-means random states were fixed before analysis and were distinct from model-training initializations. Outcomes and clinical-context variables were not used for clustering. Labels R1–R8 were assigned post hoc by increasing future-positive rate. t-SNE and UMAP were descriptive projections of the standardized probability profiles.

Six binary clinical-context domains were joined after clustering. For ordered cluster $r$ and context $d$, enrichment was

$$E_{rd} = 100\left[\Pr(C_{id} = 1 \mid R_i = r) - \Pr(C_{id} = 1)\right]. \tag{S8}$$

Printed heat-map labels show percentage-point enrichment. Colors were scaled within each domain and cannot be compared quantitatively across columns. These analyses describe output strata and post hoc context, not clinical phenotypes or treatment effects.

#### S1.8 Temporal and endpoint sensitivities

Coarse-label temporal sensitivity crossed grid resolutions of 5, 15, 30 and 60 min with horizons of 1, 2, 3 and 4 h. Internal and bidirectional source-only external AUROC and AUPRC were calculated for each model and configuration. Configuration sensitivity was the maximum minus minimum metric over the 16 settings. Refined endpoint sensitivity used 20 full-clinical runs for each endpoint, model and external direction. The source for Supplementary Fig. S6 contains 400 rows: 20 runs for five models, two endpoints and two directions.

Expanded-feature displays paired clean and expanded raw external-test estimates within model and direction; $\Delta$ was expanded minus clean. Curated-allraw displays paired curated and clean estimates generated with the same model-training seed; $\Delta$ was curated-allraw minus clean. History sensitivities refitted models after retaining both history blocks, one block or neither block. Attribution redistribution compared separately refitted XGBoost models and did not treat removed-feature SHAP mass as a causal effect. The natural-person-grouped and body-size-excluded sensitivities each reran LASSO, XGBoost, GRU, LSTM and Transformer models for 20 model-training runs in both external directions. Each sensitivity changed one factor relative to the full-clinical, `both_hist`, 60 min/4 h primary analysis; their results were compared separately with the primary means and were not interpreted as a combined factorial rerun.

#### S1.9 Calibration, target adaptation and review-policy calculations

For raw probability $\widehat{p}_{ij}$, logistic calibration estimated intercept $a$ and slope $b$ on validation predictions and applied

$$\widehat{p}_{ij}^{\mathrm{cal}} = \sigma\left[a + b\log\left(\frac{\widehat{p}_{ij}^{(\epsilon)}}{1-\widehat{p}_{ij}^{(\epsilon)}}\right)\right], \quad \widehat{p}_{ij}^{(\epsilon)} = \min\{1-\epsilon, \max(\epsilon, \widehat{p}_{ij})\}, \quad \epsilon = 10^{-6}, \tag{S9}$$

where $\sigma(v) = (1+e^{-v})^{-1}$. Test labels were not used to fit the calibration map. Isotonic and raw-probability analyses were retained as sensitivities when available.

Calibration curves were interpreted as descriptive binned observed-versus-predicted summaries. Because the persistent/recurrent future-positive label was sparse, calibration intercept and slope could be unstable in low-event strata. Non-finite or non-positive slope estimates were recorded as calibration-model failure or instability flags and were not replaced by, or interpreted as, perfect calibration. Probability claims therefore emphasized binned patterns, Brier score and log loss together with ranking, decision curves and frozen-threshold operating characteristics.

For a source-validation false-positive-rate limit $\alpha$, the fixed single threshold was

$$\tau_\alpha = \arg\max_{\tau} \mathrm{TPR}_{\mathrm{val}}(\tau) \quad \text{subject to} \quad \mathrm{FPR}_{\mathrm{val}}(\tau) \le \alpha. \tag{S10}$$

The primary limit was $\alpha = 0.05$, with 0.10 as an alert-burden sensitivity. AUPRC lift over evaluation prevalence $\pi$ was

$$L_{\mathrm{PR}} = \frac{\mathrm{AUPRC}}{\pi}. \tag{S11}$$

AUROC, AUPRC, lift, event rate, calibration and threshold operating characteristics were interpreted jointly for sparse endpoints.

Target-label adaptation was supervised and used labeled target-development ICU stays at fractions 0, 1, 5, 10, 25 and 100%. Zero was source-only validation. Figure 6 adaptation used the no-clinical-score, `both_hist`, persistent/recurrent GRU setting on a 60 min grid with a 4 h horizon. Calibration-only adaptation fixed model weights; head-only fine-tuning updated the classifier; full fine-tuning updated all trainable parameters. Subsets were sampled by ICU stay, while AUROC and AUPRC were calculated over eligible windows. Deltas were paired with the corresponding source-only results from the same model initialization and direction. Points summarized eight direction–run differences, comprising four runs in each direction; intervals were observed minima and maxima, not confidence intervals.

Decision-curve net benefit for $N$ evaluation units and threshold $\tau$ was

$$\mathrm{NB}(\tau) = \frac{\mathrm{TP}(\tau)}{N} - \frac{\mathrm{FP}(\tau)}{N}\frac{\tau}{1-\tau}. \tag{S12}$$

The Figure 6 decision curve used eligible windows as evaluation units and represents a retrospective window-review analysis.

For the final two-cutoff display, each external-test XGBoost prediction row $r$, associated with ICU stay $i$ and offset $t_{ij}$, defined

$$Z_{ijr}(\tau_{\mathrm{L}}, \tau_{\mathrm{H}}) = \begin{cases} \text{rule out}, & \widehat{p}_{ijr} \le \tau_{\mathrm{L}}, \\ \text{clinical review}, & \tau_{\mathrm{L}} < \widehat{p}_{ijr} < \tau_{\mathrm{H}}, \\ \text{alert}, & \widehat{p}_{ijr} \ge \tau_{\mathrm{H}}, \end{cases} \qquad \tau_{\mathrm{L}} < \tau_{\mathrm{H}}. \tag{S13}$$

The policy analysis used a prespecified source-only XGBoost model. Cutoffs were selected separately by direction from internal development predictions in the source database and applied unchanged to held-out external target-test predictions. The development sets contained 15,736 MIMIC-IV and 43,436 eICU prediction windows. The search targeted rule-out NPV 0.95 and alert specificity 0.95, required a low threshold no greater than 0.05 and a high threshold no less than 0.05, and returned 0.050000 and 0.783610 for MIMIC-IV-to-eICU transfer and 0.050000 and 0.702704 for eICU-to-MIMIC-IV transfer. External evaluation used 143,382 eICU and 47,082 MIMIC-IV target-test prediction rows, respectively. Main Fig. 6e pooled the two test sets only after each direction's frozen pair had been applied; pooled zone counts and rates were weighted by their observed prediction-row counts, and no common high threshold was estimated from pooled test labels. Percentile 95% intervals were calculated separately by direction from 2,000 bootstrap replicates with thresholds fixed and are reported in Supplementary Table S7 rather than on the pooled panel.

The external-test prediction table contained exactly six rows for every source–target–ICU-stay–offset key. Figure 6e reports row-level counts, but the workload analysis did not treat these rows as independent clinical alerts. For the set $\mathcal{R}_{ij}$ of prediction rows sharing stay $i$ and offset $t_{ij}$, the deduplicated clinical-window zone and outcome were

$$Z_{ij}^{\text{cw}} = \max_{r\in\mathcal{R}_{ij}} Z_{ijr}, \qquad Y_{ij}^{\text{cw}} = \max_{r\in\mathcal{R}_{ij}} Y_{ijr}, \tag{S14}$$

where the ordered zone rank was rule out < review < alert. This collapsed 190,464 prediction rows to 31,744 unique clinical windows across 4,548 ICU stays. For an 8 h shift or complete ICU stay $u$, $Z_u = \max_{(i,j)\in u} Z_{ij}^{\text{cw}}$ and $Y_u = \max_{(i,j)\in u} Y_{ij}^{\text{cw}}$. These outcomes were the maximum existing future label within the unit and were not newly adjudicated delirium episodes.

Alert episodes were constructed from deduplicated alert clinical windows within each ICU stay. The first alert began an episode; subsequent alert windows occurring before a fixed refractory interval had elapsed from that episode's start were grouped into the same episode. Eight hours was prespecified as the primary refractory interval, with 4 h and 12 h as sensitivities. Episode yield was the proportion of episodes containing at least one future-positive alert clinical window. The workload analysis involved no model training or threshold selection and reported descriptive estimates without confidence intervals. Aggregate results are in Supplementary Table S13.

An exploratory sensitivity analysis summarized each ICU stay by $S_i = \max_j \widehat{p}_{ij}^{\text{cal}}$ and selected pooled or direction-specific cutoffs in the same external-test ICU stays used for evaluation. Because cutoff selection and evaluation used the same data, the direction-specific and pooled results in Supplementary Figs. S21 and S22 are descriptive and do not support the frozen-policy result. Neither the final nor exploratory two-cutoff analyses tested prospective actionability, intervention effectiveness, workflow benefit or patient benefit.

##### S1.10 Statistical summaries and uncertainty

Assessment counts per ICU-day were compared with a two-sided, tie- and continuity-corrected asymptotic Mann–Whitney $U$ test. For model $m$, source $s$, target $t$ and coarse-label fold $k$, the paired transport difference, its mean and displayed range were

$$\begin{aligned}
\Delta_{m,k}^{s\to t} &= \text{AUROC}_{m,k}^{s\to t} - \text{AUROC}_{m,k}^{s\to s}, \\
\overline{\Delta}_m^{s\to t} &= K^{-1}\sum_{k=1}^{K} \Delta_{m,k}^{s\to t}, \\
\mathcal{R}_m^{s\to t} &= \left[\min_k \Delta_{m,k}^{s\to t}, \max_k \Delta_{m,k}^{s\to t}\right], \qquad K = 5.
\end{aligned} \tag{S15}$$

The figure reported $100\overline{\Delta}$ and $100\mathcal{R}$ in percentage points. The interval was a fold minimum–maximum range, not a confidence interval.

Primary refined analyses used 20 model-training runs per analysis cell and generally displayed individual runs, their mean and observed minimum–maximum range. Repetitions quantified training and partition sensitivity and were not independent biological replicates. No pairwise significance tests ranked the five model families. Except where stated otherwise, ICU stays rather than windows were bootstrap-resampled to preserve within-stay dependence. A sampling-uncertainty sensitivity selected one natural-person-grouped model and partition before resampling target ICU stays 2,000 times separately for each model and transfer direction, retaining all windows from each selected stay. Its percentile 95% intervals were conditional on that model and partition and did not replace the 20-run ranges. The final frozen-threshold prediction-row analysis used 2,000 bootstrap replicates with thresholds held fixed; the deduplicated workload analysis was descriptive and did not use confidence intervals. The exploratory same-test-selected ICU-stay analysis used 1,000 stay-level resamples with cutoffs fixed; because selection was not repeated and had already used the test cohort, those intervals quantify conditional sampling variation only. All tests were two sided; no multiplicity adjustment was applied to descriptive or prespecified sensitivity panels.

#### S2 Supplementary Results

##### S2.1 Temporal sensitivity of the coarse weak-label benchmark

Internal coarse-label discrimination was comparatively stable at 15–60 min resolutions, whereas the 5 min setting reduced performance for several MIMIC-IV models (Supplementary Fig. S1). External results were more configuration dependent and direction asymmetric (Supplementary Fig. S2). AUPRC showed a corresponding pattern internally and externally (Supplementary Figs. S3 and S4). Across the 16 temporal configurations, sequence models generally had larger max–min ranges than tabular models, particularly under transfer (Supplementary Fig. S5). These panels support the use of a fixed temporal specification for primary refined comparisons and caution against selecting a resolution or horizon after inspecting external test results.

#### S2.2 Refined endpoint and probability-profile sensitivities

The primary full-clinical endpoint sensitivity included 20 model-training runs per analysis cell. Models for persistence or recurrence retained substantial external discrimination, whereas mean AUROCs for strict new onset ranged from 0.50 to 0.58 (Supplementary Fig. S6). Probability-profile analyses produced post hoc risk gradients across model families (Supplementary Fig. S7). For each model, the context heat map, t-SNE projection, UMAP projection, cluster sizes and future-positive rates were generated from the same ICU-stay-cluster assignment table (Supplementary Figs. S8–S11). Thus, the displayed projection legends and context annotations refer to the same R1–R8 assignments; the projections remain descriptive and do not establish discrete clinical phenotypes.

#### S2.3 Feature expansion and history dependence

Broader feature representations produced heterogeneous rather than uniform changes in external performance. With expanded features, GRU AUROC increased in both transfer directions ($\Delta = +0.015$ and $+0.017$), whereas Transformer AUROC decreased in MIMIC-IV-to-eICU transfer ($\Delta = -0.022$) and was essentially unchanged in the reverse direction (+0.002; Supplementary Fig. S12). AUPRC increased for LSTM in both directions (+0.038 and +0.024) but decreased for GRU and Transformer in both directions (Supplementary Fig. S13). Curated-allraw augmentation left AUROC nearly unchanged for XGBoost and the three sequence models (absolute $\Delta \leq 0.003$) but reduced LASSO AUROC by 0.089 and 0.062 in the two transfer directions (Supplementary Fig. S14). Curated-allraw AUPRC changes were also model and direction dependent, ranging from $-0.044$ for LASSO in eICU-to-MIMIC-IV transfer to +0.012 for LSTM in MIMIC-IV-to-eICU transfer (Supplementary Fig. S15). Thus, neither additional representation produced a consistent external-performance gain across models, metrics and transfer directions.

Removal of all assessment history produced the largest AUROC reduction in every external and internal model comparison (Supplementary Figs. S16 and S17). AUPRC lift generally decreased with history removal, although the after-24 h-only variant occasionally matched or slightly exceeded the both-history value for an individual model. The XGBoost redistribution analysis showed that removal of history shifted mean absolute SHAP attribution toward neurologic or consciousness, observation-process, laboratory and other non-history domains (Supplementary Fig. S18). These changes describe redistribution within refitted models and do not identify causal importance.

#### S2.4 Partition and body-size integrity sensitivities

Natural-person grouping retained the principal direction asymmetry and above-chance external discrimination for every model (Supplementary Table S8). Across the ten model–direction cells, mean AUROC changed by $-0.002$ to 0.021 and mean AUPRC by $-0.004$ to 0.016 relative to the primary ICU-stay-grouped analysis. Absolute person-grouped AUROC ranged from 0.865 to 0.946 for MIMIC-IV-to-eICU transfer and from 0.770 to 0.904 for eICU-to-MIMIC-IV transfer. The separate body-size-excluded rerun changed mean AUROC by $-0.004$ to 0.020 and mean AUPRC by $-0.003$ to 0.006. These comparisons used 20 model-training runs for each of five models and two transfer directions. Because one sensitivity changed the grouping unit and the other changed three features, their deltas were each calculated against the primary analysis and not against one another.

A separate bootstrap analysis quantified target-cohort sampling uncertainty (Supplementary Table S12). One natural-person-grouped model and partition were selected before target ICU stays were sampled with replacement in 2,000 replicates, preserving all windows within each sampled stay. The resulting percentile intervals were calculated for each of five model families and both transfer directions. They quantify sampling uncertainty conditional on that model and partition and should be read alongside, not in place of, the 20-run minimum–maximum ranges.

#### S2.5 Transfer adaptation and retrospective review-policy sensitivities

The AUROC sensitivity analysis reproduced the data-size dependence observed for AUPRC (Supplementary Fig. S19). Mean within-run paired AUROC changes for full fine-tuning were negative at the 1% target fraction, positive at 5–25%, and largest at 100% target-data use. Head-only fine-tuning showed smaller mean changes, while monotonic recalibration left AUROC unchanged. Intervals in this panel are observed minimum–maximum ranges across eight direction–run results, comprising four model-training runs in each transfer direction, and are not confidence intervals.

Calibration transportability was also direction dependent (Supplementary Fig. S20). Internal and external calibration curves did not follow a common probability mapping, and the two external directions showed opposite departures in their upper risk bins. Because events were sparse, calibration slopes or intercepts that were non-finite or had a non-positive slope were treated as estimation-instability flags rather than perfect calibration. These window-level binned curves support target-specific calibration assessment but do not establish a deployed review policy.

The final two-cutoff analysis used direction-specific XGBoost cutoffs selected only in source/internal development predictions and applied unchanged to external target-test prediction rows (main Fig. 6e and Supplementary Table S7). MIMIC-IV-to-eICU cutoffs of 5.0% and 78.4% allocated 91.3%, 8.4% and 0.3% of prediction rows to rule out, review and alert, with future-positive rates of 0.18%, 2.84% and 13.78%. eICU-to-MIMIC-IV cutoffs of 5.0% and 70.3% allocated 88.7%, 9.2% and 2.0%, with rates of 1.51%, 7.01% and 12.24%. Main Fig. 6e pools the resulting assignments only for presentation; its 190,464-row denominator was not used to select a common cutoff. Row-level alert capture was 9.2% and 11.0%.

Because each source–target–ICU-stay–offset combination occurred in six prediction rows, the workload analysis collapsed these rows to 31,744 clinical windows and 4,548 ICU stays before counting alerts (Supplementary Table S13). Clinical-window alert fractions were 1.1% for MIMIC-IV-to-eICU and 7.3% for eICU-to-MIMIC-IV transfer; the corresponding ICU-stay fractions were 4.9% and 12.2%. Across 8 h shifts, 1.9% and 8.2% were assigned to alert. Under the primary 8 h refractory rule, 141 and 452 alert episodes remained, corresponding to 8.1 and 16.1 episodes per 100 ICU stays; future-positive episode yields were 15.6% and 14.6%. The pooled workload comprised 827 alert clinical windows, 427 ICU stays with at least one alert and 593 alert episodes. Four-hour and 12 h refractory sensitivities are also reported in Supplementary Table S13. Direction-specific and pooled ICU-stay sensitivity analyses selected and evaluated cutoffs in the same external test data (Supplementary Figs. S21 and S22) and are therefore reported as exploratory only. Neither the frozen policy nor the workload analysis evaluated prospective actionability, an intervention, workflow benefit or patient benefit.

### S3 Supplementary Tables

Table S1: Cohort construction and refined-endpoint inventory. Counts are reported as ICU stays / windows / endpoint-positive windows. Positive-window counts apply only to endpoint-eligibility rows, and denominators are ICU-stay based.

| Analysis stage | eICU | MIMIC-IV |
|---|---|---|
| Complete hourly grid | 28,321 / 2,496,432 / – | 49,828 / 5,881,343 / – |
| Post-24 h grid | 24,060 / 1,845,924 / – | 47,181 / 4,694,559 / – |
| Direct-assessment cells, complete grid | 27,216 / 380,780 / – | 49,828 / 457,471 / – |
| Direct-assessment cells, post-24 h | 20,387 / 261,345 / – | 42,425 / 333,261 / – |
| Strict new-onset eligible | 9,131 / 117,519 / 1,067 | 9,823 / 26,405 / 854 |
| Persistent/recurrent eligible | 11,572 / 157,056 / 782 | 18,736 / 52,530 / 1,224 |

Table S2: Database-specific delirium-assessment mapping. Raw-row counts describe the candidate mapping before hourly collapse and are not independent assessments or ICU stays.

| Source | Raw-value rule | Harmonized state | Raw rows |
|---|---|---|---|
| eICU charted candidates | Positive, present, yes, 1 or true | Explicit positive | 44,982 |
| eICU charted candidates | Negative, absent, no, 0 or false | Explicit negative | 459,043 |
| eICU charted candidates | Unable, UTA, not assessed, cannot assess, not applicable or unknown | Unable to assess | 30,806 |

| Source | Raw-value rule | Harmonized state | Raw rows |
|---|---|---|---|
| eICU charted candidates | Header-like or unmapped candidate value | No mapped evidence | 293,266 |
| eICU ICDSC numeric | Valid 0–8 score; positive if score $\geq 4$, otherwise negative | Explicit positive or negative | Included above |
| MIMIC-IV item 228332 | Mapped positive values | Explicit positive | 118,632 |
| MIMIC-IV item 228332 | Mapped negative values | Explicit negative | 358,477 |
| MIMIC-IV item 228332 | Mapped unable-to-assess values | Unable to assess | 129,796 |
| MIMIC-IV item 229326 | Mapped positive values | Explicit positive | 134,249 |
| MIMIC-IV item 229326 | Mapped negative values | Explicit negative | 207,012 |
| MIMIC-IV RASS item 228096 | RASS $\leq -4$ with no direct positive or negative evidence | Unable to assess | Not separately tabulated |
| Within-cell conflict rule | Simultaneous positive and negative evidence | Flagged conflict; positive point evidence | Not separately tabulated |

Table S3: Feature-construction settings used for the primary anchor tables. The schema-validated per-column dictionary is provided as Supplementary Data 1.

| Component | Verified setting |
|---|---|
| Full-clinical representation | 137 feature columns in the primary persistent/recurrent `both_hist` anchor inventory. |
| No-clinical-score representation | 124 feature columns after removal of RASS-, GCS-, pain-score-, APACHE-, OASIS- and related score fields. |
| Body-size-excluded representation | 134 full-clinical features after removing only weight, height and body mass index. |
| Numeric measurements | Most recent value, observed-within-window indicator and elapsed time since observation. |
| Lookback windows | 6 h for vital, respiratory and neurologic measurements; 24 h for laboratory measurements; 6 h for medication, vasoactive, respiratory-support and process-event indicators unless otherwise specified. |
| History variants | `no_hist`, `only_1st_hist`, `only_after_hist` and `both_hist`; the primary persistent/recurrent setting used `both_hist`. |
| History summaries | Counts, indicators and recency of direct, explicit-positive, explicit-negative and unable-to-assess states, calculated at or before the anchor. |
| No prior post-24 h direct assessment | Recency filled with 168 h and paired with a binary indicator denoting absence of a prior post-24 h direct assessment. |

Table S4: Model-training settings used for the primary refined analyses.

| Model or component | Setting |
|---|---|
| LASSO | Class-balanced $L_1$-penalized logistic regression; `saga` solver; maximum 3,000 iterations. |
| XGBoost | Binary logistic loss; histogram tree construction; 500 estimators; maximum depth 3; learning rate 0.03; subsample 0.85; column subsample 0.85; minimum child weight 5; $L_2$ regularization 2.0; source-training positive-class weighting. |
| GRU | One recurrent layer; hidden dimension 96; dropout 0.25. |
| LSTM | One recurrent layer; hidden dimension 96; dropout 0.25; recurrent state concatenated with the current-anchor dense representation before classification. |
| Transformer | One causal self-attention layer; embedding dimension 96; four heads; feed-forward dimension 256; dropout 0.25; sinusoidal positional encoding. |
| GCS/RASS score references | Descriptive only. eICU used current score plus a within-stay first difference and pre-fold analysis-table mean imputation; MIMIC-IV used current score only and training-fold median imputation. Both used training-fold scaling, class-balanced logistic regression and fivefold ICU-stay-stratified cross-validation. |
| Neural optimization | Batch size 16; maximum sequence length 512; AdamW learning rate $10^{-3}$; weight decay $10^{-4}$; weighted binary cross-entropy; positive-class weight capped at 100; gradient-norm clipping at 5.0. |
| Source training | Maximum 30 epochs; minimum 6 epochs before early stopping; patience 5. |
| Target fine-tuning | Maximum 20 epochs. |
| Coarse-label repetition | Fivefold ICU-stay-stratified cross-validation; folds are held-out partitions, not model-training runs. |

| Model or component | Setting |
|---|---|
| Primary refined repetition | 20 model-training runs per analysis cell, each initialized with a distinct random seed. |
| Target-label adaptation repetition | Four model-training runs per transfer direction, yielding eight direction–run results at each target-data fraction. |

Table S5: Calibration, threshold-selection and uncertainty settings. The final two-cutoff policy used development-selected thresholds applied unchanged to external test prediction rows; same-test-selected ICU-stay results are reported separately as exploratory sensitivity analyses.

| Component | Setting |
|---|---|
| Database split | Primary 70% training, 15% validation and 15% test split grouped by ICU stay; natural-person grouping was evaluated separately in Supplementary Table S8. |
| Logistic calibration | Platt-type map fit on source-validation predictions and applied unchanged to held-out test predictions; probability clipping $\epsilon = 10^{-6}$. |
| Sparse calibration rule | Binned curves are descriptive. Non-finite or non-positive calibration slopes are failure or instability flags, not perfect calibration. |
| Single-threshold rule | Maximize source-validation recall subject to false-positive rate $\leq 0.05$; false-positive rate $\leq 0.10$ retained as a burden sensitivity analysis. |
| Final two-cutoff model and unit | Prespecified source-only XGBoost; external-test prediction rows for Fig. 6e and deduplicated clinical windows for workload. |
| Threshold-selection data | Source-database internal development predictions: 15,736 MIMIC-IV windows and 43,436 eICU windows. |
| Selection targets | Rule-out NPV 0.95 and alert specificity 0.95; low threshold ceiling 0.05 and high threshold floor 0.05. |
| Frozen external evaluation | Direction-specific cutoffs applied unchanged to 143,382 eICU and 47,082 MIMIC-IV target-test prediction rows. |
| Workload collapse | Six prediction rows per source–target–ICU-stay–offset combination; 190,464 rows collapsed to 31,744 clinical windows, 14,753 observed 8 h shifts and 4,548 ICU stays. |
| Alert episodes | Fixed refractory interval from episode onset; 8 h primary, with 4 h and 12 h sensitivities. |
| Frozen-policy intervals | Percentile 95% intervals from 2,000 bootstrap replicates with thresholds held fixed. |
| Exploratory ICU-stay analysis | Maximum calibrated window score per stay; cutoffs selected and evaluated in the same external test cohort; 1,000 ICU-stay bootstrap resamples with cutoffs fixed. |

Table S6: Supervised target-label adaptation settings represented in Figure 6.

| Component | Setting |
|---|---|
| Task | Persistent/recurrent delirium; 60 min grid; 4 h horizon; no-clinical-score; `both_hist`; GRU. |
| Labeled target fractions | 0, 1, 5, 10, 25 and 100% of target-development ICU stays; 0% denotes source-only validation. |
| Strategies | Calibration only, head-only fine-tuning and full fine-tuning; target-scratch and other tabular adaptations were sensitivity references where available. |
| Sampling and evaluation units | Adaptation subsets sampled by ICU stay with stay-level outcome stratification; AUROC and AUPRC calculated over eligible prediction windows. |
| Pairing | Each adapted estimate paired with the corresponding source-only estimate from the same model initialization and transfer direction. |
| Current repetitions and intervals | Four model-training runs in each of two transfer directions; displayed intervals are the observed minimum–maximum range across eight direction–run results, not confidence intervals. |

Table S7: Development-selected frozen two-cutoff prediction-row operating summaries. Cutoffs were selected from source-database internal development predictions and applied unchanged to external target-test prediction rows. Deduplicated workload is reported separately in Supplementary Table S13.

| Metric | MIMIC-IV-to-eICU | eICU-to-MIMIC-IV |
|---|---|---|
| Development windows / prevalence | 15,736 / 2.49% | 43,436 / 0.42% |
| External test prediction rows / future positive | 143,382 / 630 | 47,082 / 1,050 |
| Low cutoff, $\tau_{\mathrm{L}}$ | 5.00% | 5.00% |
| High cutoff, $\tau_{\mathrm{H}}$ | 78.36% | 70.27% |
| Rule-out prediction rows, $n$ (%) | 130,918 (91.31%) | 41,783 (88.75%) |
| Rule-out future-positive rate | 0.18% | 1.51% |
| Rule-out NPV | 99.82% | 98.49% |
| Review prediction rows, $n$ (%) | 12,043 (8.40%) | 4,351 (9.24%) |
| Review future-positive rate | 2.84% | 7.01% |
| Alert prediction rows, $n$ (%) | 421 (0.29%) | 948 (2.01%) |
| Alert future-positive rate / PPV | 13.78% | 12.24% |
| Alert specificity | 99.75% | 98.19% |
| Alert sensitivity | 9.21% | 11.05% |
| Selection / evaluation | MIMIC-IV internal development / eICU external test | eICU internal development / MIMIC-IV external test |

Table S8: External persistent/recurrent performance in the primary analysis and two independent integrity sensitivities. Values are mean AUROC / mean AUPRC across 20 model-training runs. Parentheses give sensitivity minus primary. Body-size exclusion and natural-person grouping each changed one factor and were not combined.

| Model | Direction | Primary | Body-size excluded (Δ) | Natural-person grouped (Δ) |
|---|---|---|---|---|
| LASSO | MIMIC-IV→eICU | 0.867 / 0.051 | 0.867 / 0.053 (+0.0002 / +0.0021) | 0.865 / 0.057 (−0.0012 / +0.0060) |
| XGBoost | MIMIC-IV→eICU | 0.944 / 0.098 | 0.944 / 0.098 (+0.0001 / +0.0007) | 0.946 / 0.108 (+0.0013 / +0.0106) |
| GRU | MIMIC-IV→eICU | 0.882 / 0.044 | 0.885 / 0.045 (+0.0025 / +0.0015) | 0.888 / 0.047 (+0.0057 / +0.0033) |
| LSTM | MIMIC-IV→eICU | 0.867 / 0.043 | 0.880 / 0.048 (+0.0130 / +0.0056) | 0.887 / 0.059 (+0.0205 / +0.0157) |
| Transformer | MIMIC-IV→eICU | 0.857 / 0.055 | 0.874 / 0.053 (+0.0163 / −0.0020) | 0.869 / 0.053 (+0.0120 / −0.0018) |
| LASSO | eICU→MIMIC-IV | 0.764 / 0.061 | 0.760 / 0.060 (−0.0036 / −0.0013) | 0.770 / 0.061 (+0.0063 / +0.0000) |
| XGBoost | eICU→MIMIC-IV | 0.900 / 0.148 | 0.899 / 0.146 (−0.0003 / −0.0024) | 0.904 / 0.153 (+0.0045 / +0.0045) |
| GRU | eICU→MIMIC-IV | 0.762 / 0.079 | 0.770 / 0.077 (+0.0083 / −0.0022) | 0.773 / 0.085 (+0.0108 / +0.0055) |
| LSTM | eICU→MIMIC-IV | 0.756 / 0.076 | 0.777 / 0.082 (+0.0202 / +0.0050) | 0.774 / 0.078 (+0.0173 / +0.0019) |
| Transformer | eICU→MIMIC-IV | 0.773 / 0.086 | 0.770 / 0.083 (−0.0026 / −0.0029) | 0.771 / 0.082 (−0.0023 / −0.0037) |

Table S9: Analysis-grid size and delirium-label availability. Counts are reported at the ICU-stay or temporal-grid-window level, as indicated. Percentages for label states use all windows in the corresponding database as denominator. The final row reports the positive percentage and the denominator of windows with an unambiguous explicit positive or negative assessment. This table was moved from the main text to satisfy the six-display-item Article format.

| Characteristic | eICU | MIMIC-IV |
|---|---|---|
| ICU stays | 28,321 | 49,828 |
| Temporal-grid windows | 2,496,432 | 5,881,343 |
| No assessment record, $n$ (%) | 2,099,176 (84.1) | 5,288,613 (89.9) |
| Unable to assess, $n$ (%) | 16,476 (0.7) | 135,259 (2.3) |
| Assessed label state, $n$ (%) | 380,780 (15.3) | 457,471 (7.8) |
| Explicit positive among unambiguous assessments, % (denominator $n$) | 9.5 (380,518) | 29.6 (429,016) |

Table S10: Coarse-label model performance. Ranges span mean performance across LASSO, XGBoost, GRU, LSTM and Transformer models in the primary 60 min-grid, 4 h-horizon, full-clinical setting. AUPRC lift is AUPRC divided by endpoint prevalence in the evaluation database. Internal estimates are means across five folds; external estimates use the corresponding five-fold ensembles. This table was moved from the main text to satisfy the six-display-item Article format.

| Training database | Test database | AUROC range | AUPRC-lift range |
|---|---|---|---|
| eICU | eICU | 0.88–0.90 | 4.5–5.2 |
| MIMIC-IV | MIMIC-IV | 0.87–0.92 | 2.3–2.7 |
| eICU | MIMIC-IV | 0.78–0.83 | 1.8–2.0 |
| MIMIC-IV | eICU | 0.66–0.77 | 1.7–2.5 |

Table S11: ICU-stay characteristics of the persistent/recurrent analysis cohort. Continuous variables are median (interquartile range), and categorical variables are count (percentage). Characteristics were collapsed once per ICU stay from the full-clinical, both-history eligible-anchor tables. ICU-type indicators are not mutually exclusive. Race or ethnicity, mortality and complete ICU length of stay were not present in these harmonized anchor tables and are therefore not reported.

| Characteristic | eICU | MIMIC-IV |
|---|---|---|
| ICU stays, n | 11,572 | 18,736 |
| Age, years, median (IQR) | 66.0 (54.0–77.0) | 65.0 (53.0–75.0) |
| Male sex, n (%) | 6,087 (52.6) | 10,560 (56.4) |
| Medical ICU indicator, n (%) | 1,164 (10.1) | 7,478 (39.9) |
| Surgical ICU indicator, n (%) | 6,289 (54.3) | 6,670 (35.6) |
| Cardiac/coronary ICU indicator, n (%) | 2,050 (17.7) | 4,170 (22.3) |
| Neurological ICU indicator, n (%) | 1,473 (12.7) | 2,977 (15.9) |
| Other or unspecified ICU type, n (%) | 596 (5.2) | 21 (0.1) |
| First eligible anchor, h, median (IQR) | 27.0 (25.0–42.0) | 38.0 (28.0–57.0) |
| Eligible windows per stay, median (IQR) | 5 (2–13) | 2 (1–3) |
| Direct assessment in first 24 h, n (%) | 9,974 (86.2) | 16,490 (88.0) |
| Positive assessment in first 24 h, n (%) | 1,704 (14.7) | 6,024 (32.2) |
| Any future persistent/recurrent-positive window, n (%) | 500 (4.3) | 1,200 (6.4) |

Table S12: ICU-stay-clustered sampling uncertainty for one prespecified natural-person-grouped model and partition. Points are window-level external AUROC and AUPRC; parentheses are percentile 95% intervals from 2,000 bootstrap resamples of target ICU stays, retaining all windows from each sampled stay. These intervals quantify target-cohort sampling uncertainty conditional on that model and partition; they complement, rather than replace, the 20-run minimum–maximum ranges that quantify training and split sensitivity.

| Model | Direction | ICU stays | AUROC (95% CI) | AUPRC (95% CI) |
|---|---|---|---|---|
| LASSO | MIMIC-IV→eICU | 1,731 | 0.846 (0.796–0.888) | 0.043 (0.020–0.093) |
| XGBoost | MIMIC-IV→eICU | 1,731 | 0.943 (0.925–0.958) | 0.110 (0.071–0.176) |
| GRU | MIMIC-IV→eICU | 1,731 | 0.851 (0.809–0.887) | 0.030 (0.019–0.047) |
| LSTM | MIMIC-IV→eICU | 1,731 | 0.886 (0.849–0.915) | 0.042 (0.026–0.071) |
| Transformer | MIMIC-IV→eICU | 1,731 | 0.851 (0.808–0.888) | 0.049 (0.026–0.096) |
| LASSO | eICU→MIMIC-IV | 2,785 | 0.731 (0.683–0.774) | 0.073 (0.057–0.095) |
| XGBoost | eICU→MIMIC-IV | 2,785 | 0.905 (0.891–0.918) | 0.155 (0.123–0.202) |
| GRU | eICU→MIMIC-IV | 2,785 | 0.799 (0.768–0.826) | 0.087 (0.066–0.117) |
| LSTM | eICU→MIMIC-IV | 2,785 | 0.761 (0.726–0.794) | 0.080 (0.059–0.113) |
| Transformer | eICU→MIMIC-IV | 2,785 | 0.754 (0.713–0.795) | 0.089 (0.068–0.124) |

Table S13: Frozen-policy workload and event-yield analysis after deduplication of external-test prediction rows. The Figure 6e data contained six prediction rows per source–target–ICU-stay–offset combination. For workload estimation, rows at the same combination were collapsed to one clinical window using the highest assigned risk zone. Shift and ICU-stay zones were the maximum clinical-window zone within that unit; their outcomes were the maximum existing future label and were not independently adjudicated episodes. The 8 h refractory analysis was primary; 4 h and 12 h were sensitivity analyses. Pooled values were calculated only after direction-specific frozen thresholds had been applied.

| Metric | MIMIC-IV→eICU | eICU→MIMIC-IV | Pooled |
|---|---|---|---|
| External-test prediction rows, $n$ | 143,382 | 47,082 | 190,464 |
| Unique clinical windows, $n$ | 23,897 | 7,847 | 31,744 |
| Clinical windows assigned to review or alert, $n$ (%) | 5,147 (21.5%) | 2,407 (30.7%) | 7,554 (23.8%) |
| Alert clinical windows, $n$ (%) | 258 (1.1%) | 569 (7.3%) | 827 (2.6%) |
| Future-positive clinical windows in alert zone, $n/N$ (%) | 34/105 (32.4%) | 67/175 (38.3%) | 101/280 (36.1%) |
| Observed 8 h ICU-stay shifts, $n$ | 8,742 | 6,011 | 14,753 |
| 8 h shifts assigned to review or alert, $n$ (%) | 1,987 (22.7%) | 1,895 (31.5%) | 3,882 (26.3%) |
| Alert 8 h shifts, $n$ (%) | 164 (1.9%) | 495 (8.2%) | 659 (4.5%) |
| Future-positive 8 h shifts in alert zone, $n/N$ (%) | 25/70 (35.7%) | 71/172 (41.3%) | 96/242 (39.7%) |
| External-test ICU stays, $n$ | 1,737 | 2,811 | 4,548 |
| ICU stays assigned to review or alert, $n$ (%) | 426 (24.5%) | 959 (34.1%) | 1,385 (30.5%) |
| ICU stays with at least one alert, $n$ (%) | 85 (4.9%) | 342 (12.2%) | 427 (9.4%) |
| Future-positive ICU stays with an alert, $n/N$ (%) | 29/66 (43.9%) | 84/170 (49.4%) | 113/236 (47.9%) |
| Alert episodes, 4 h refractory, $n$ (per 100 stays) | 197 (11.3 per 100 stays) | 554 (19.7 per 100 stays) | 751 (16.5 per 100 stays) |
| Future-positive alert episodes, 4 h refractory, $n/N$ (%) | 23/197 (11.7%) | 66/554 (11.9%) | 89/751 (11.9%) |
| Alert episodes, 8 h refractory, $n$ (per 100 stays) | 141 (8.1 per 100 stays) | 452 (16.1 per 100 stays) | 593 (13.0 per 100 stays) |
| Future-positive alert episodes, 8 h refractory, $n/N$ (%) | 22/141 (15.6%) | 66/452 (14.6%) | 88/593 (14.8%) |
| Alert episodes, 12 h refractory, $n$ (per 100 stays) | 125 (7.2 per 100 stays) | 417 (14.8 per 100 stays) | 542 (11.9 per 100 stays) |
| Future-positive alert episodes, 12 h refractory, $n/N$ (%) | 23/125 (18.4%) | 66/417 (15.8%) | 89/542 (16.4%) |

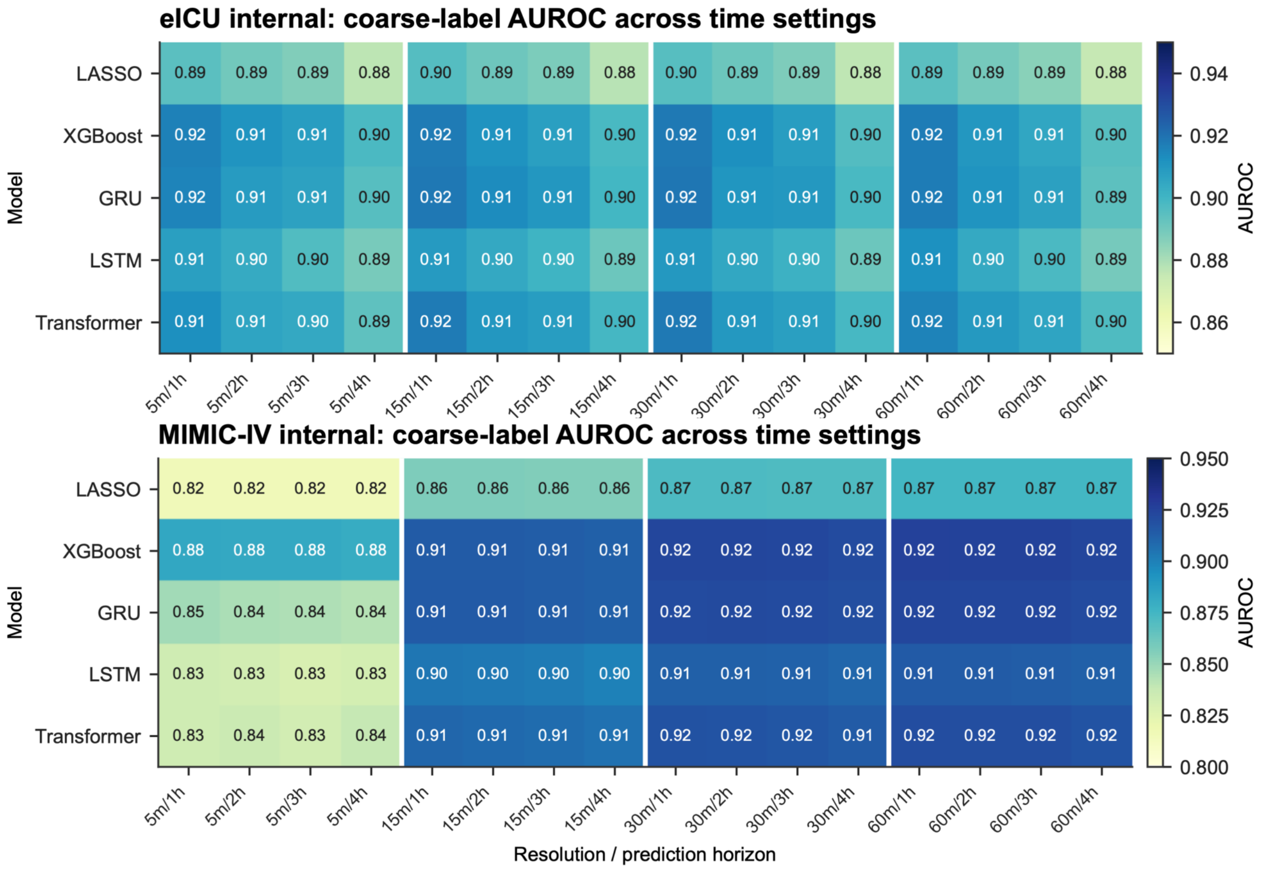


Figure S1: Internal coarse-label AUROC across temporal settings. Heat maps show eICU (top) and MIMIC-IV (bottom) internal AUROC for five model families across the Cartesian product of grid resolution (5, 15, 30 or 60 min) and prediction horizon (1–4 h). Cell labels are AUROC values. White vertical lines separate grid resolutions.

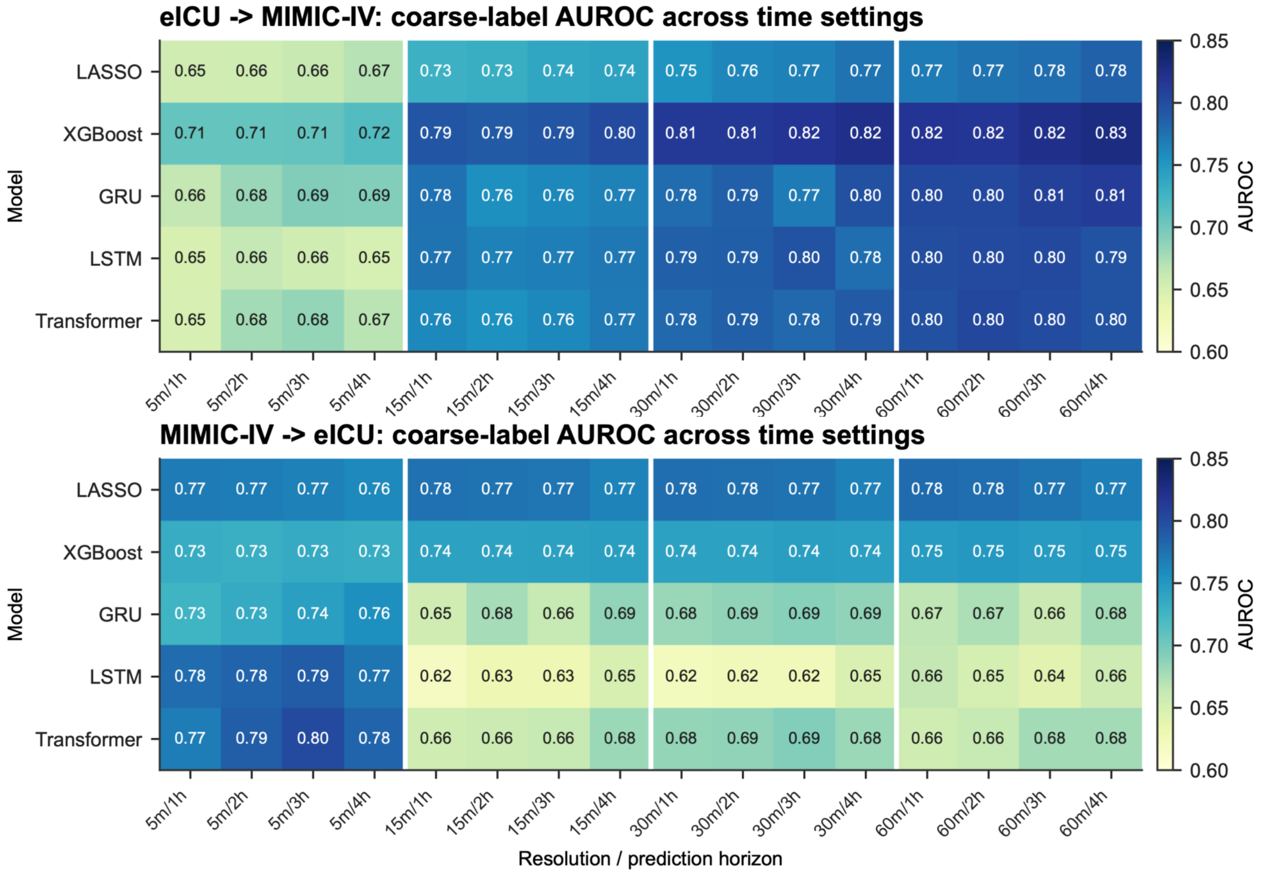


Figure S2: Source-only external coarse-label AUROC across temporal settings. Heat maps show eICU-to-MIMIC-IV (top) and MIMIC-IV-to-eICU (bottom) transfer for the same 16 grid-resolution and prediction-horizon combinations. Models were applied to the target database without target-database retraining.

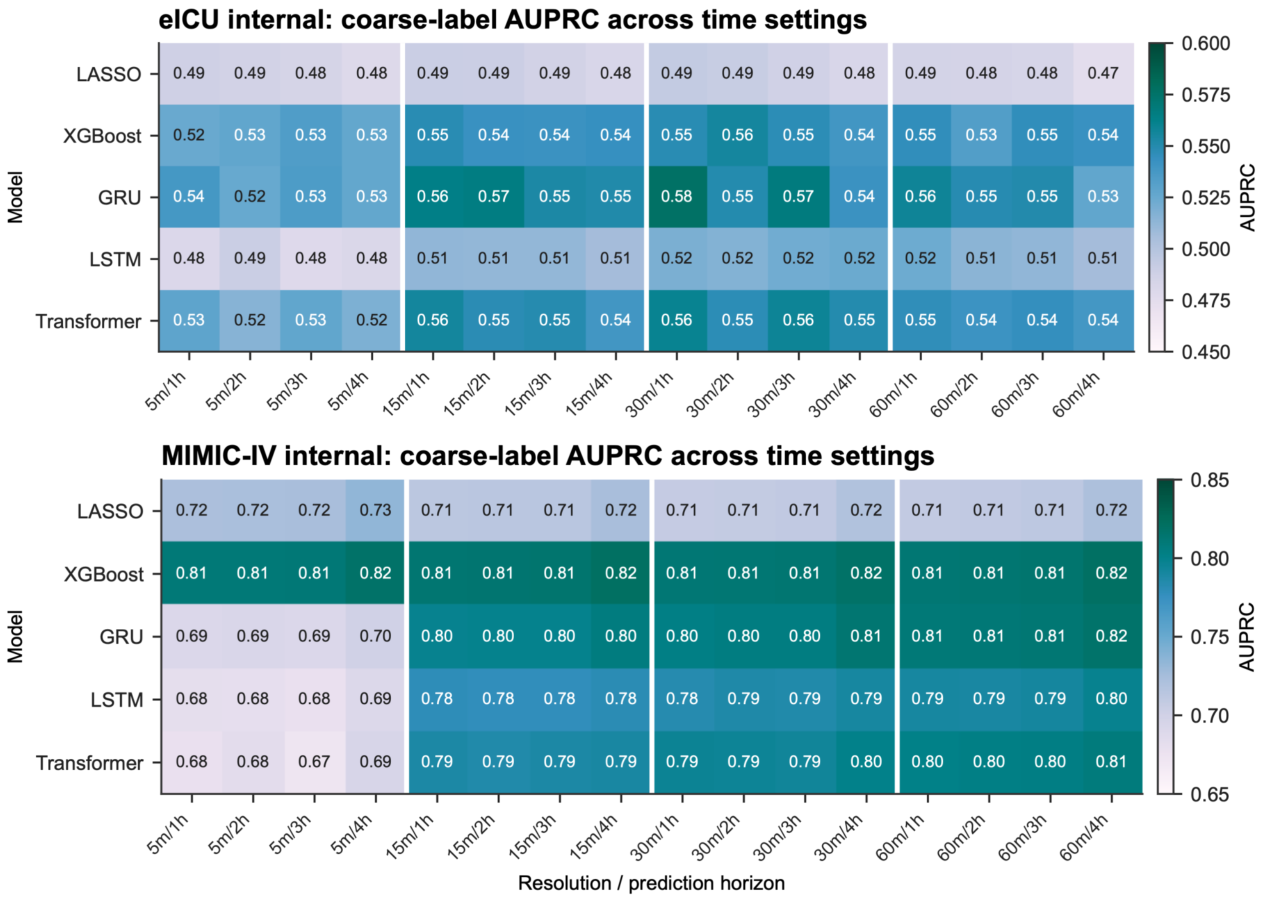


Figure S3: Internal coarse-label AUPRC across temporal settings. Heat maps show eICU (top) and MIMIC-IV (bottom) AUPRC for five model families across four grid resolutions and four prediction horizons. Because AUPRC depends on prevalence, these absolute values should be interpreted within the corresponding database and endpoint.

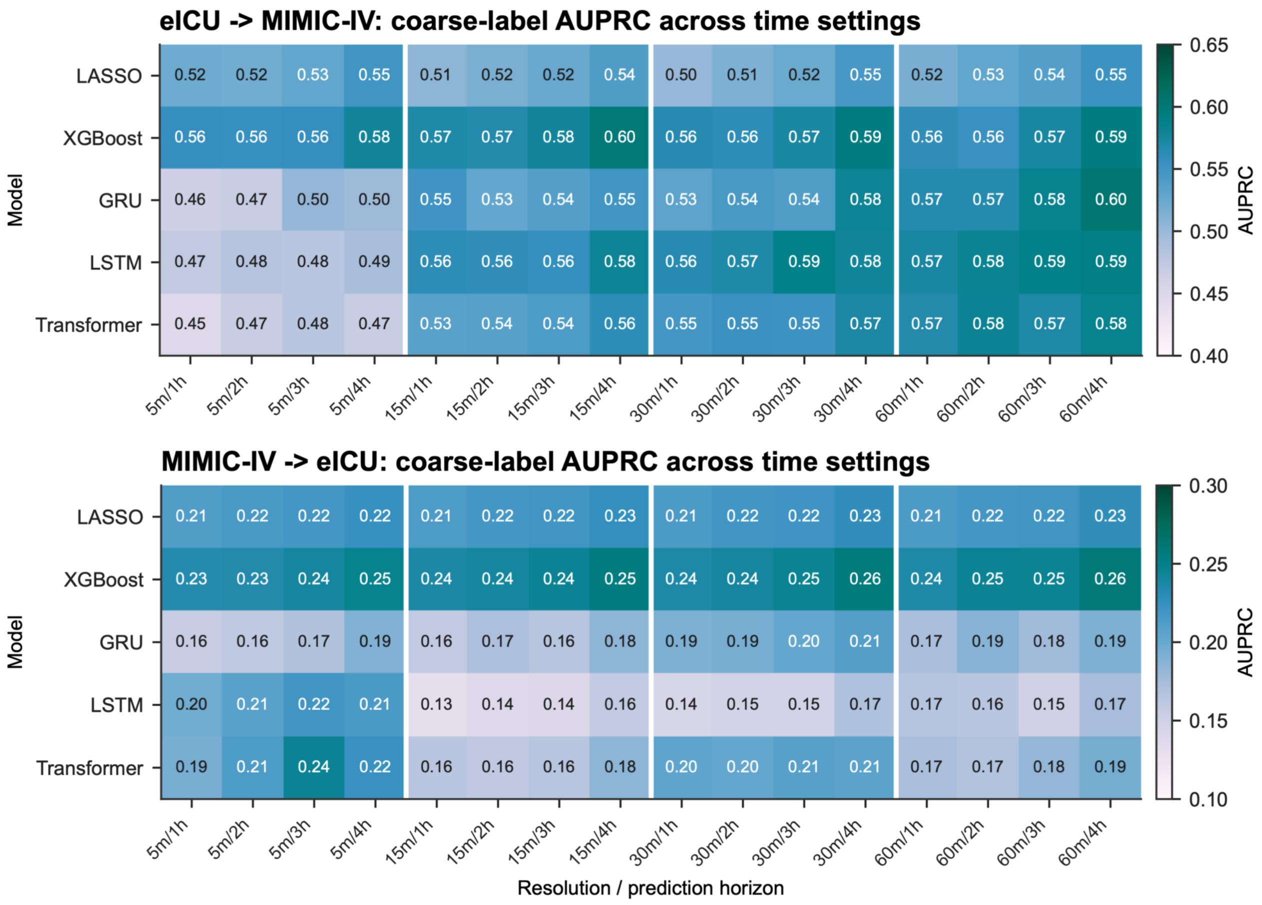


Figure S4: Source-only external coarse-label AUPRC across temporal settings. Heat maps show eICU-to-MIMIC-IV (top) and MIMIC-IV-to-eICU (bottom) results for the same 16 temporal configurations. The database-specific color scales reflect different endpoint prevalences and should not be compared by color alone.

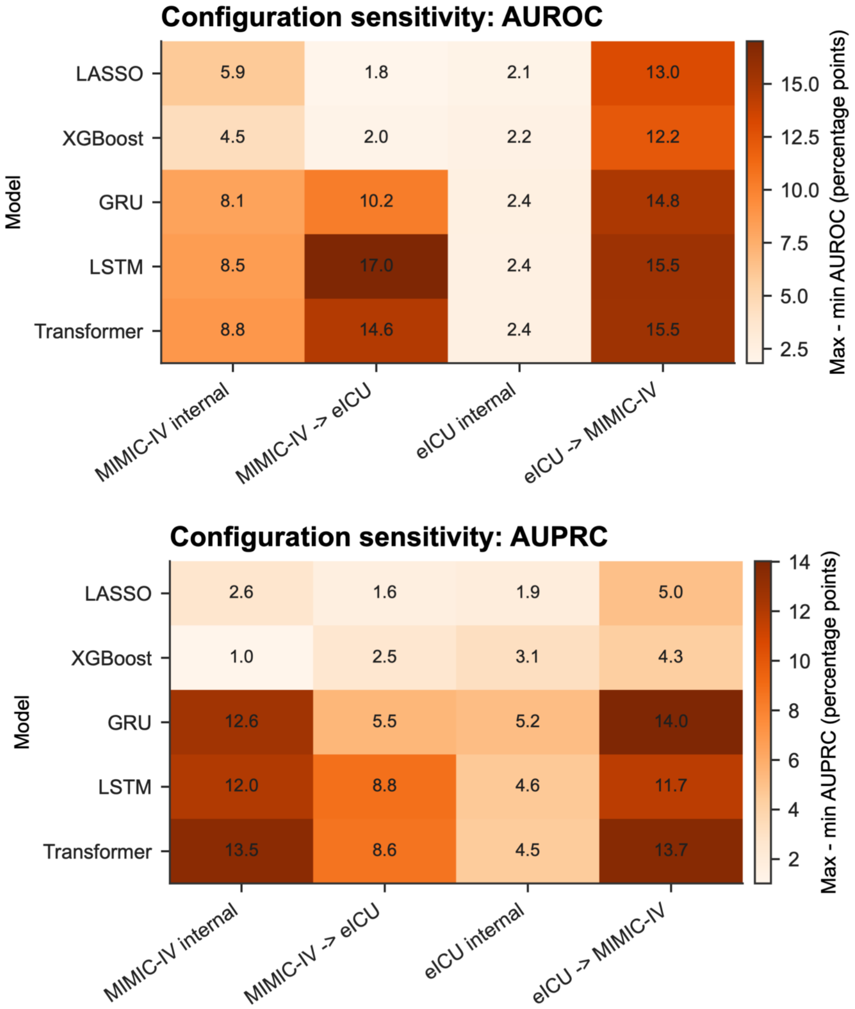


Figure S5: Sensitivity to temporal configuration. The displayed value is the maximum minus minimum performance over the 16 resolution–horizon configurations, in percentage points, for AUROC (top) and AUPRC (bottom). Columns distinguish MIMIC-IV internal, MIMIC-IV-to-eICU, eICU internal and eICU-to-MIMIC-IV evaluation. Larger values indicate greater sensitivity to temporal specification, not uncertainty intervals.

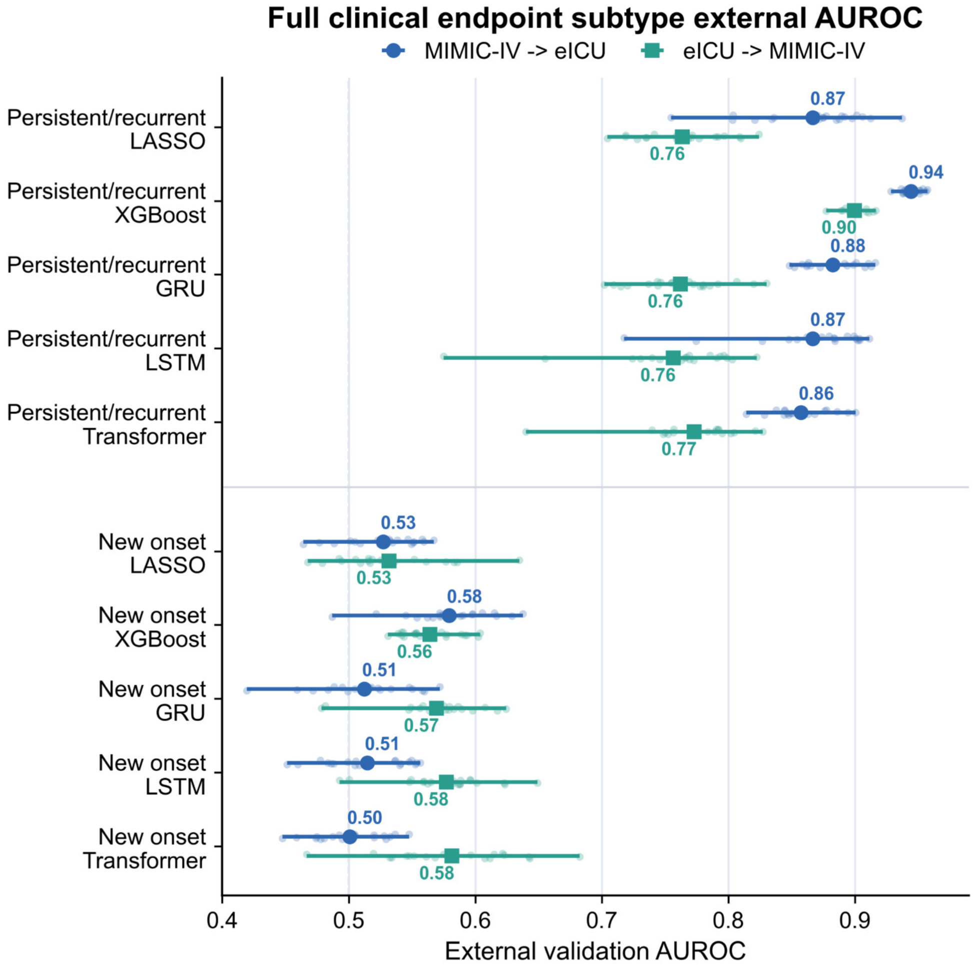


Figure S6: External AUROC by refined endpoint subtype in the full-clinical feature view. Persistent/recurrent and strict new-onset endpoints are shown for MIMIC-IV-to-eICU (blue) and eICU-to-MIMIC-IV (green) transfer. Small points denote 20 model-training runs for each model, endpoint and direction; large symbols denote their mean and horizontal intervals denote the minimum–maximum range across runs. The two endpoints have different eligibility masks and estimands, so their performance difference is not an ablation of the same outcome.

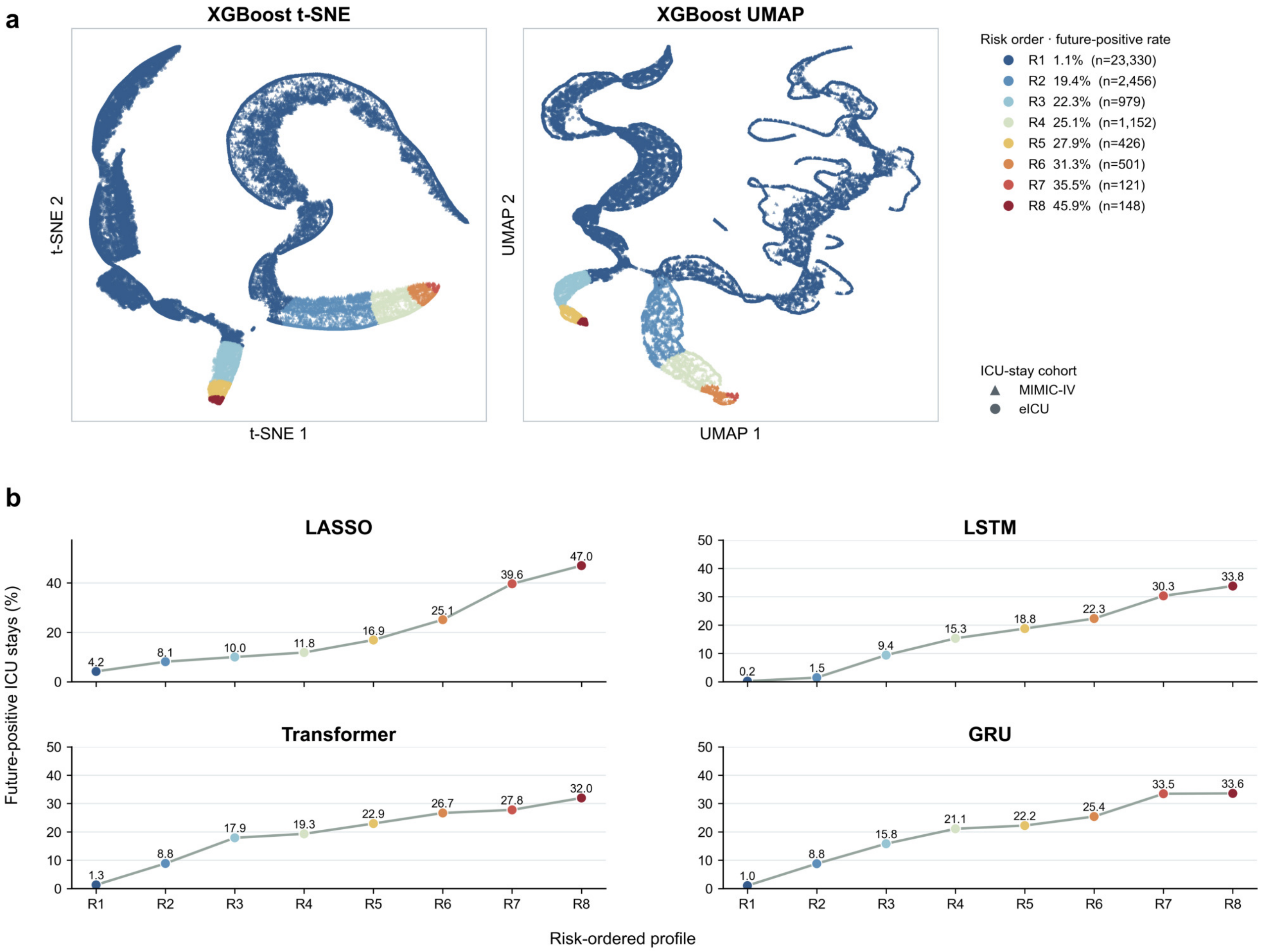


Figure S7: ICU-stay probability-profile projections and model-specific risk gradients. **a**, XGBoost-specific t-SNE and UMAP projections of standardized ICU-stay probability profiles. Color denotes the eight K-means clusters ordered post hoc by future-positive rate; marker shape denotes the database cohort. **b**, ICU-stay-level future-positive rate across risk-ordered profiles for LASSO, LSTM, Transformer and GRU. Cluster sizes, displayed rates and coordinates were derived from the same model-specific assignment data. Outcomes were used to order and annotate clusters after fitting, not as clustering inputs. Projection geometry is descriptive and does not establish discrete clinical phenotypes.

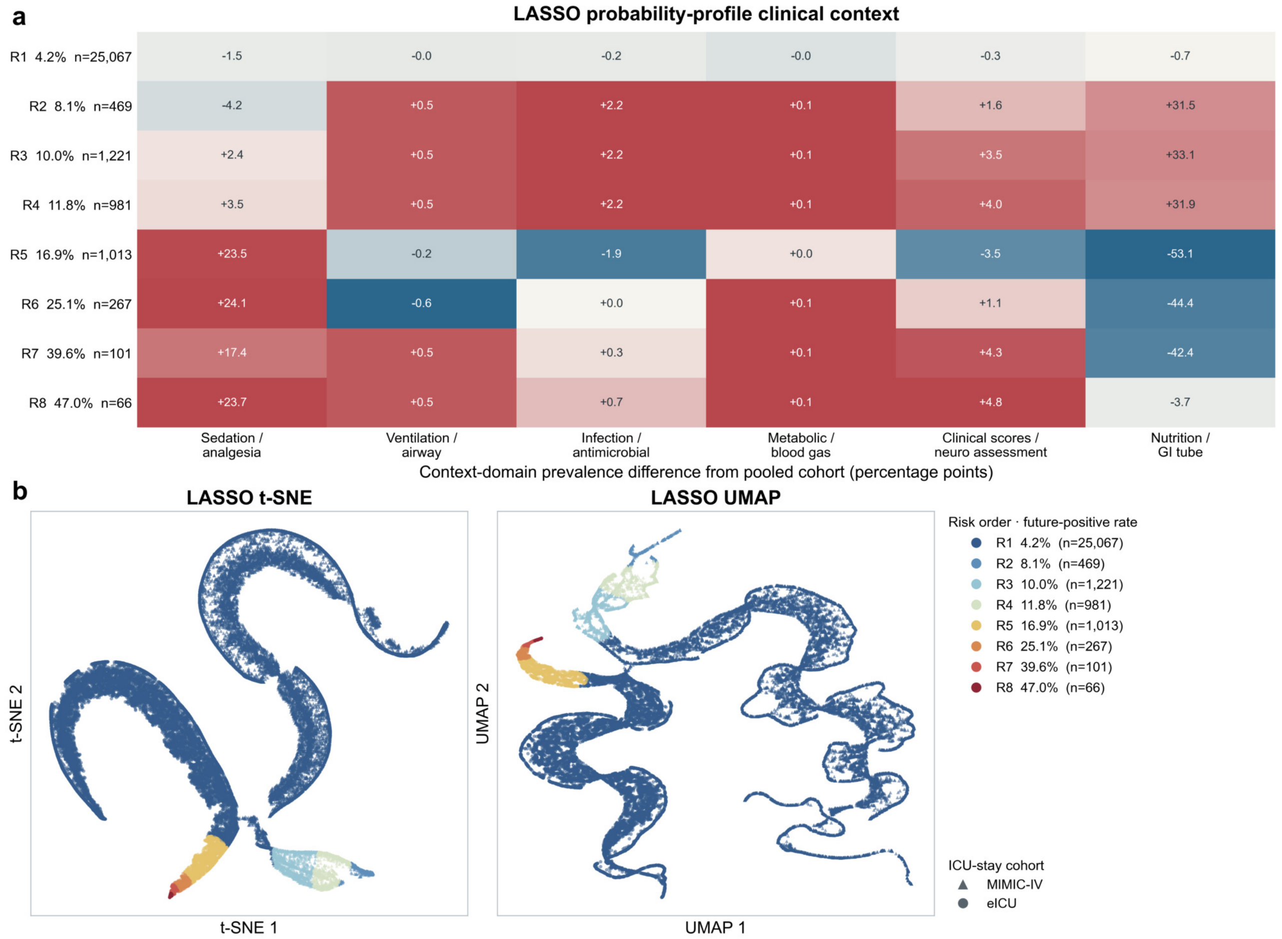


Figure S8: LASSO probability-profile context and projections. **a**, Difference between each cluster's context-domain prevalence and the pooled domain prevalence, in percentage points. Printed values are unscaled differences; color is scaled within each domain and cannot be compared quantitatively across columns. **b**, LASSO-specific t-SNE and UMAP projections colored by post hoc risk order, with shapes denoting the ICU-stay cohort. Both panels, cluster sizes and displayed future-positive rates derive from the same ICU-stay-cluster assignment. Context variables and outcomes were not used to fit K-means clusters.

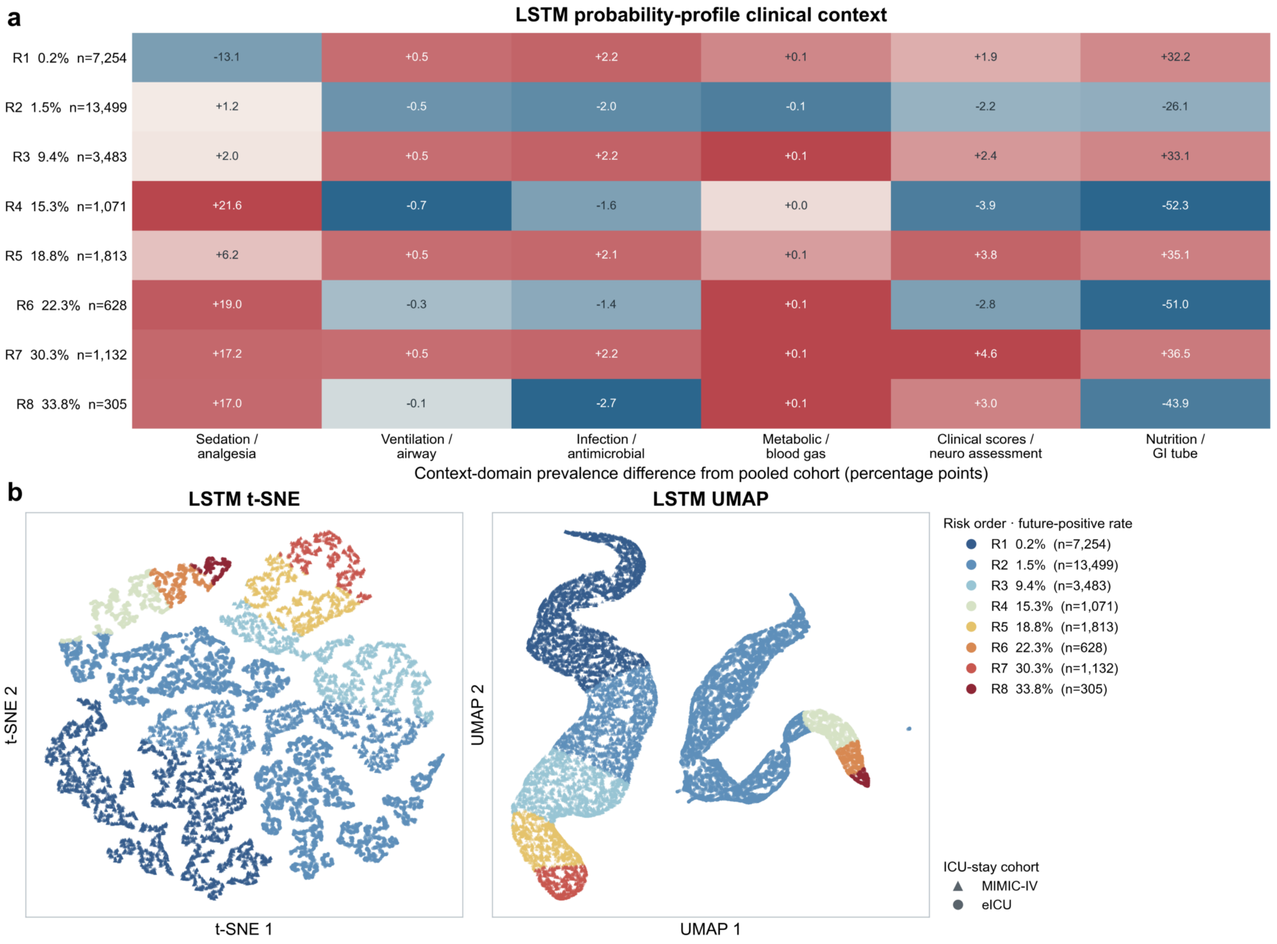


Figure S9: LSTM probability-profile context and projections. Panel definitions follow Supplementary Fig. S8. The context heat map, t-SNE projection, UMAP projection, cluster sizes and displayed future-positive rates derive from the same ICU-stay-cluster assignment.

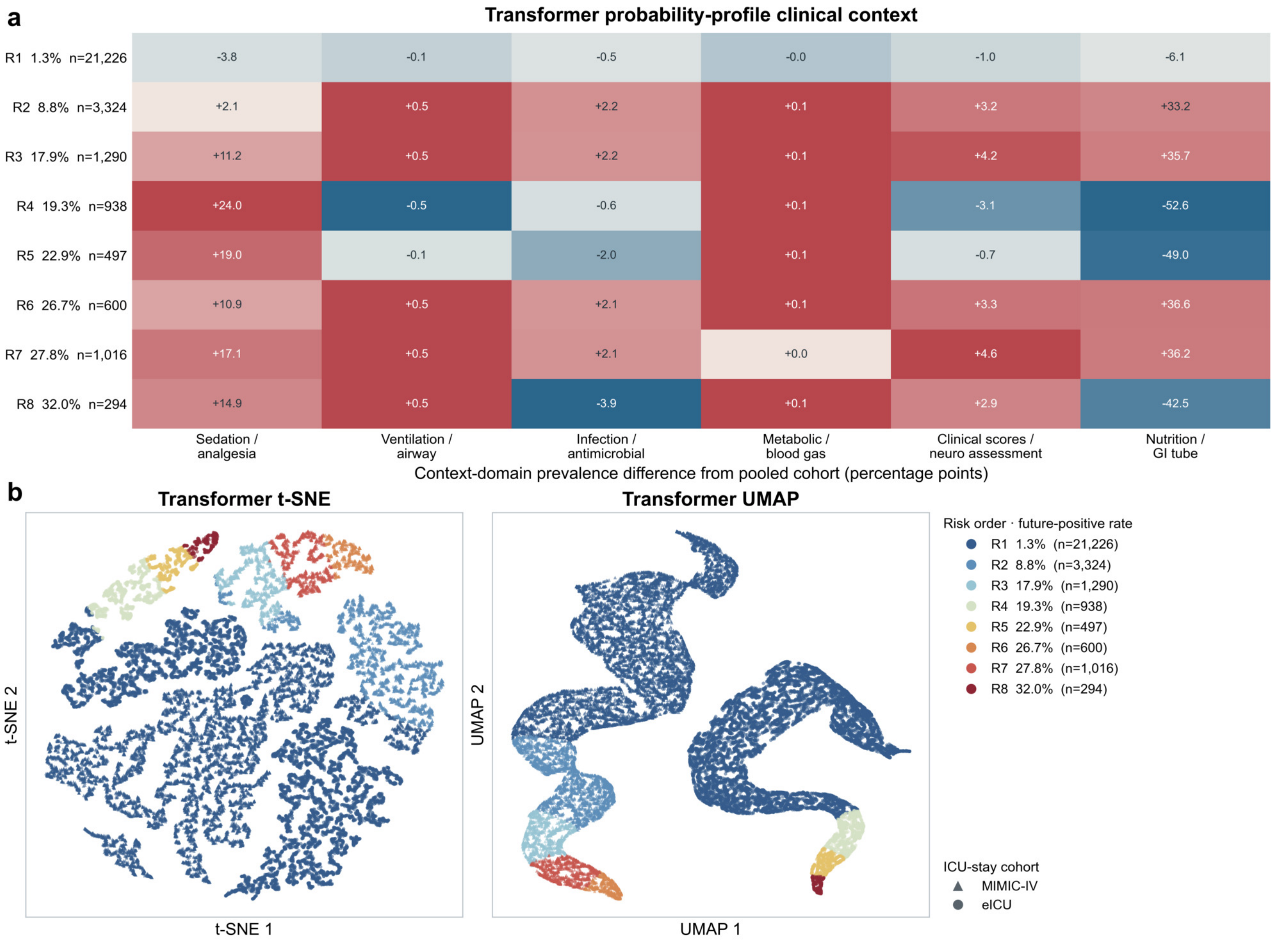


Figure S10: Transformer probability-profile context and projections. Panel definitions follow Supplementary Fig. S8. The context heat map, t-SNE projection, UMAP projection, cluster sizes and displayed future-positive rates derive from the same ICU-stay-cluster assignment.

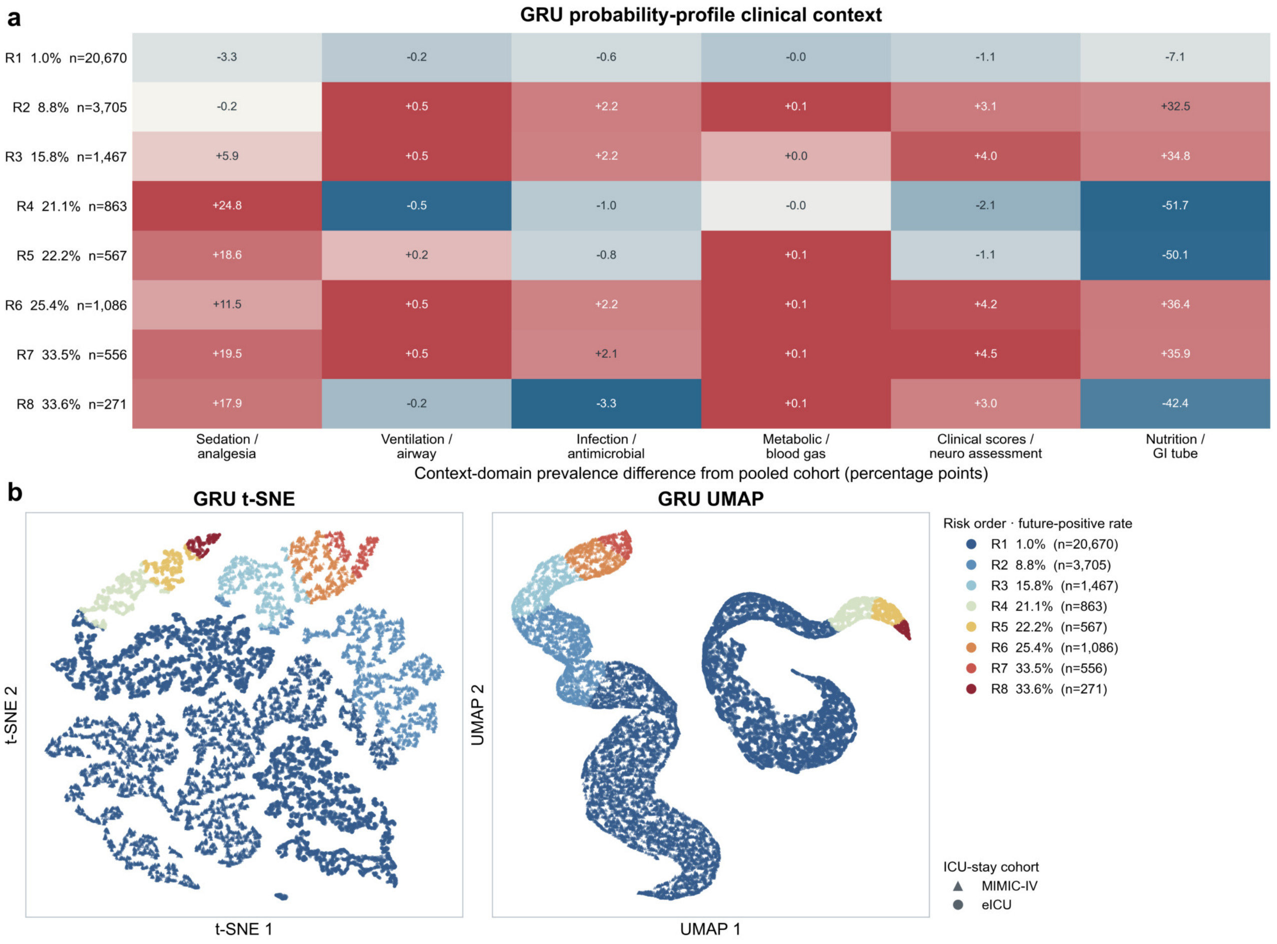


Figure S11: GRU probability-profile context and projections. Panel definitions follow Supplementary Fig. S8. The context heat map, t-SNE projection, UMAP projection, cluster sizes and displayed future-positive rates derive from the same ICU-stay-cluster assignment.

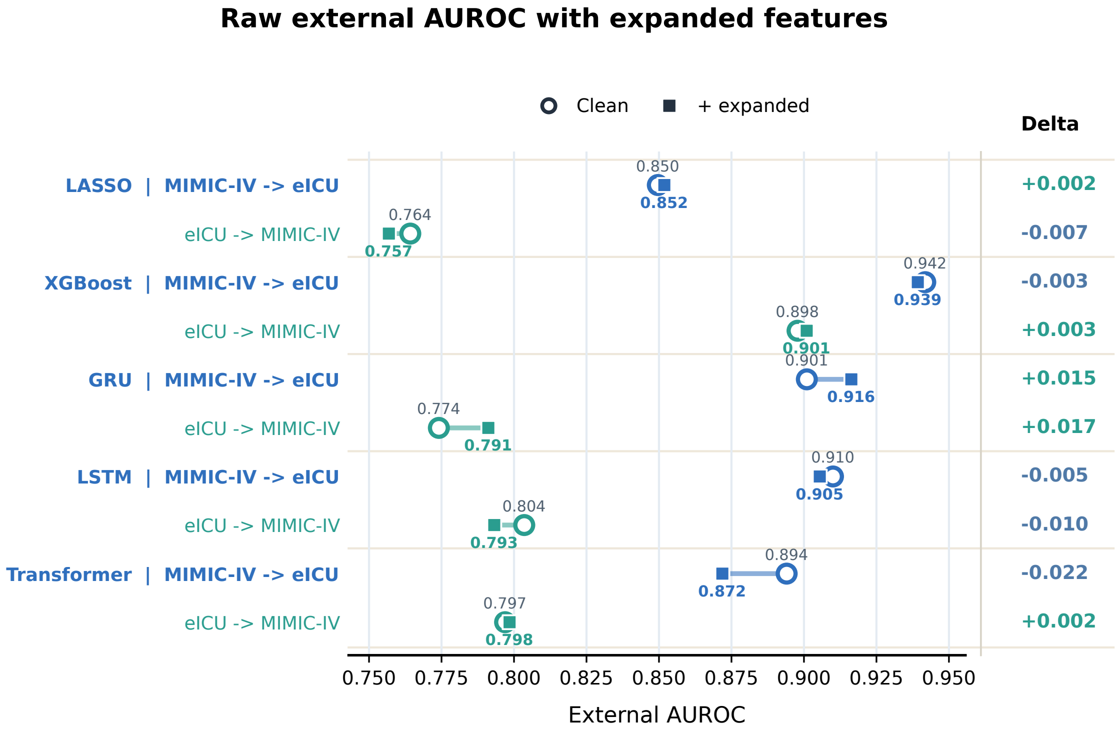


Figure S12: Raw external AUROC after expanded feature augmentation. Hollow circles show estimates from the clean prespecified feature representation, filled squares show estimates from the expanded representation, and horizontal segments connect the two estimates within each model and transfer direction. The right-hand $\Delta$ is expanded minus clean; positive values favor the expanded representation. The display contains point estimates and does not show uncertainty intervals.

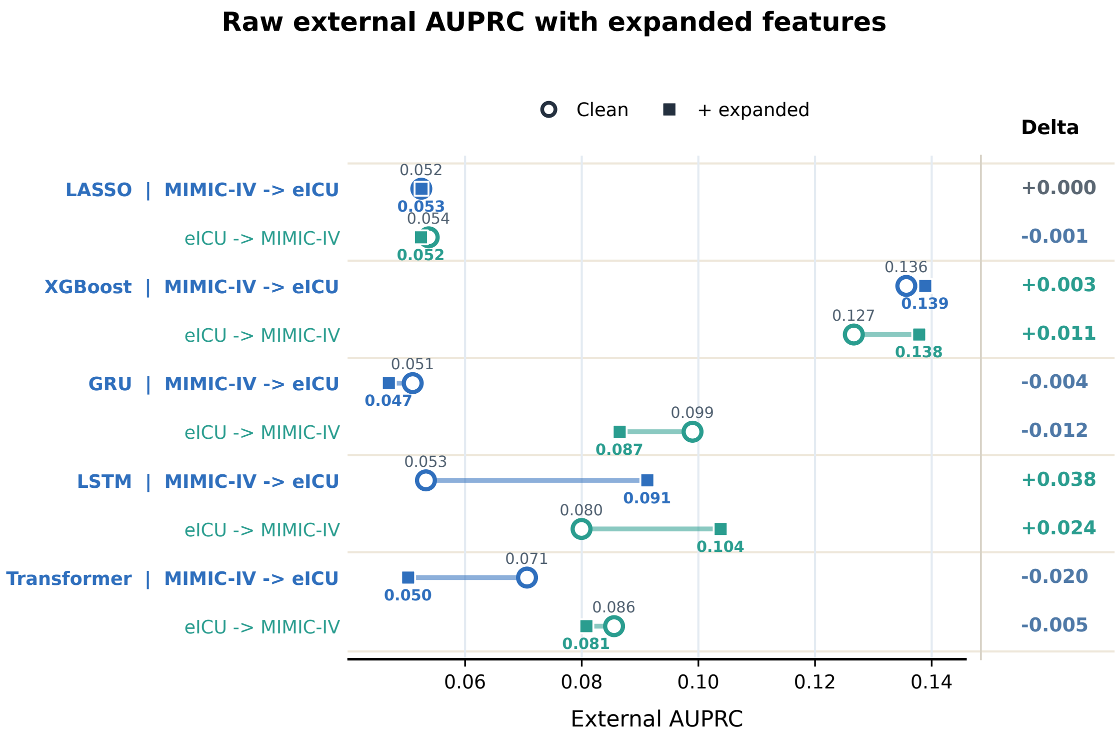


Figure S13: Raw external AUPRC after expanded feature augmentation. Hollow circles show estimates from the clean prespecified feature representation, filled squares show estimates from the expanded representation, and horizontal segments connect the two estimates within each model and transfer direction. The right-hand $\Delta$ is expanded minus clean; positive values favor the expanded representation. Because AUPRC depends on outcome prevalence, absolute values should be interpreted within each target database. The display contains point estimates and does not show uncertainty intervals.

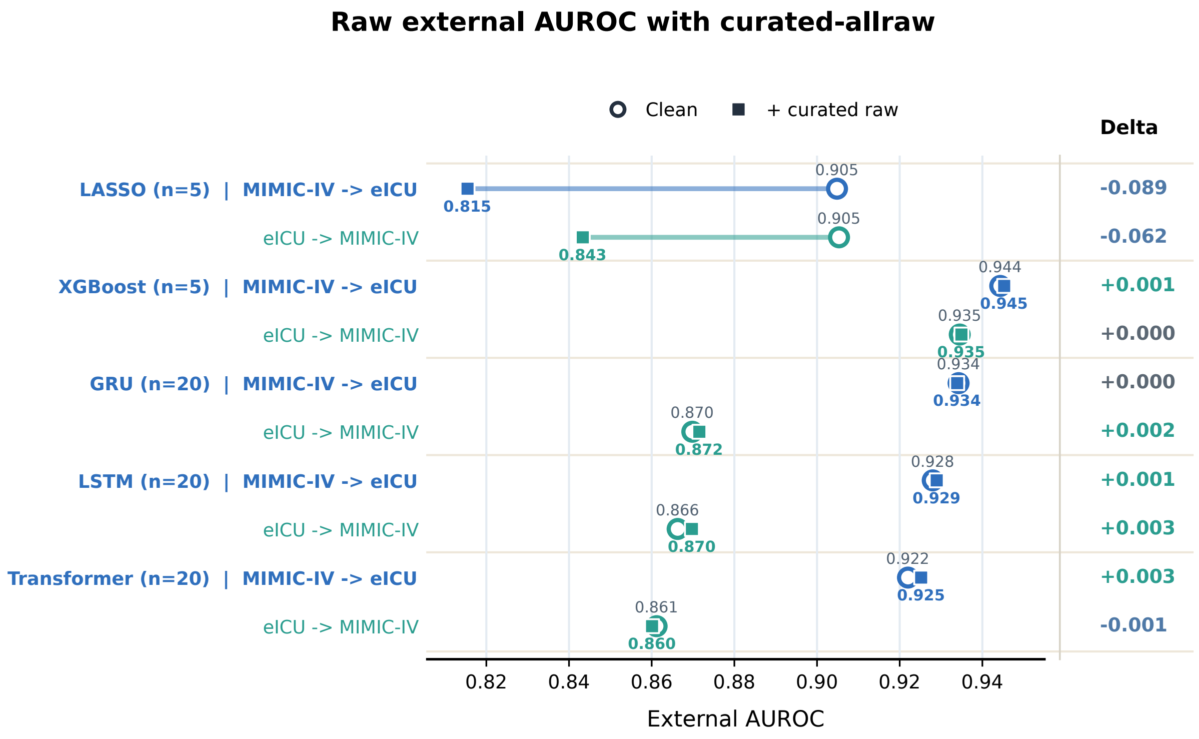


Figure S14: Raw external AUROC after curated-allraw feature augmentation. Hollow circles show mean estimates from the clean prespecified feature representation, filled squares show mean estimates from the curated-allraw representation, and horizontal segments connect the paired means within each model and transfer direction. The right-hand $\Delta$ is curated-allraw minus clean; positive values favor curated-allraw. Tabular models used five paired model-training runs and sequence models used 20. Connecting segments are paired changes, not uncertainty intervals.

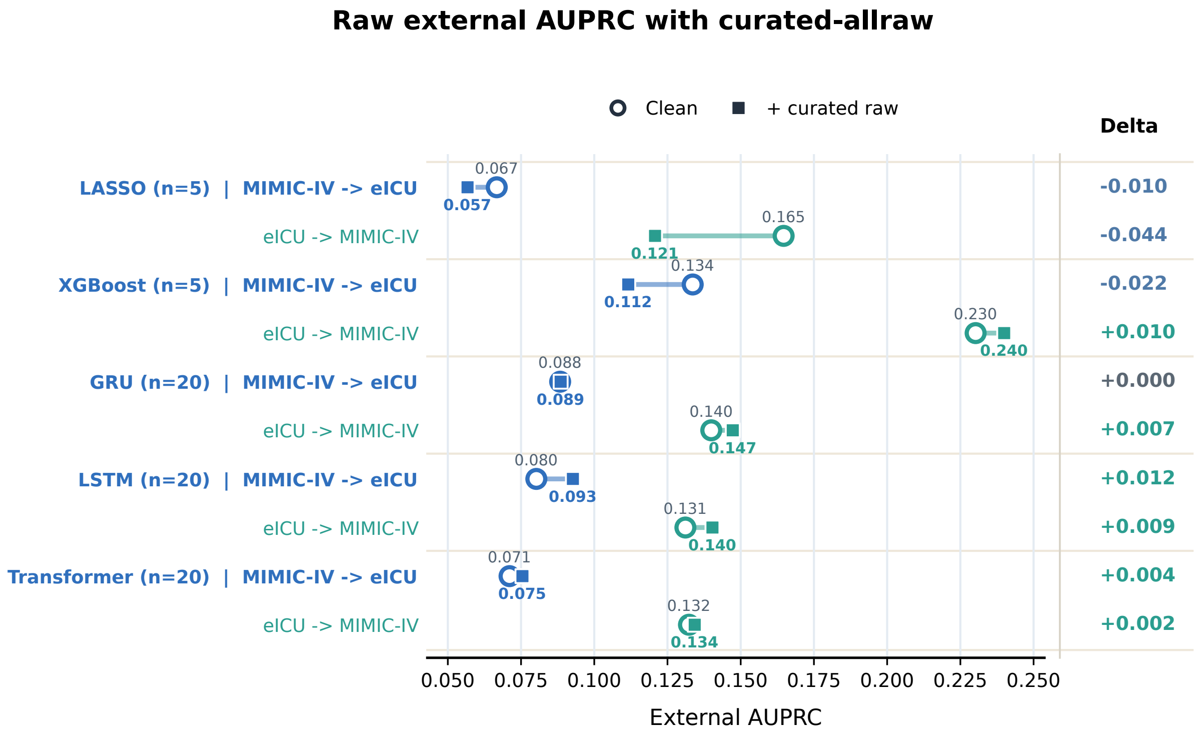


Figure S15: Raw external AUPRC after curated-allraw feature augmentation. Hollow circles show mean estimates from the clean prespecified feature representation, filled squares show mean estimates from the curated-allraw representation, and horizontal segments connect the paired means within each model and transfer direction. The right-hand $\Delta$ is curated-allraw minus clean; positive values favor curated-allraw. Tabular models used five paired model-training runs and sequence models used 20. Because AUPRC depends on outcome prevalence, absolute values should be interpreted within each target database. Connecting segments are paired changes, not uncertainty intervals.

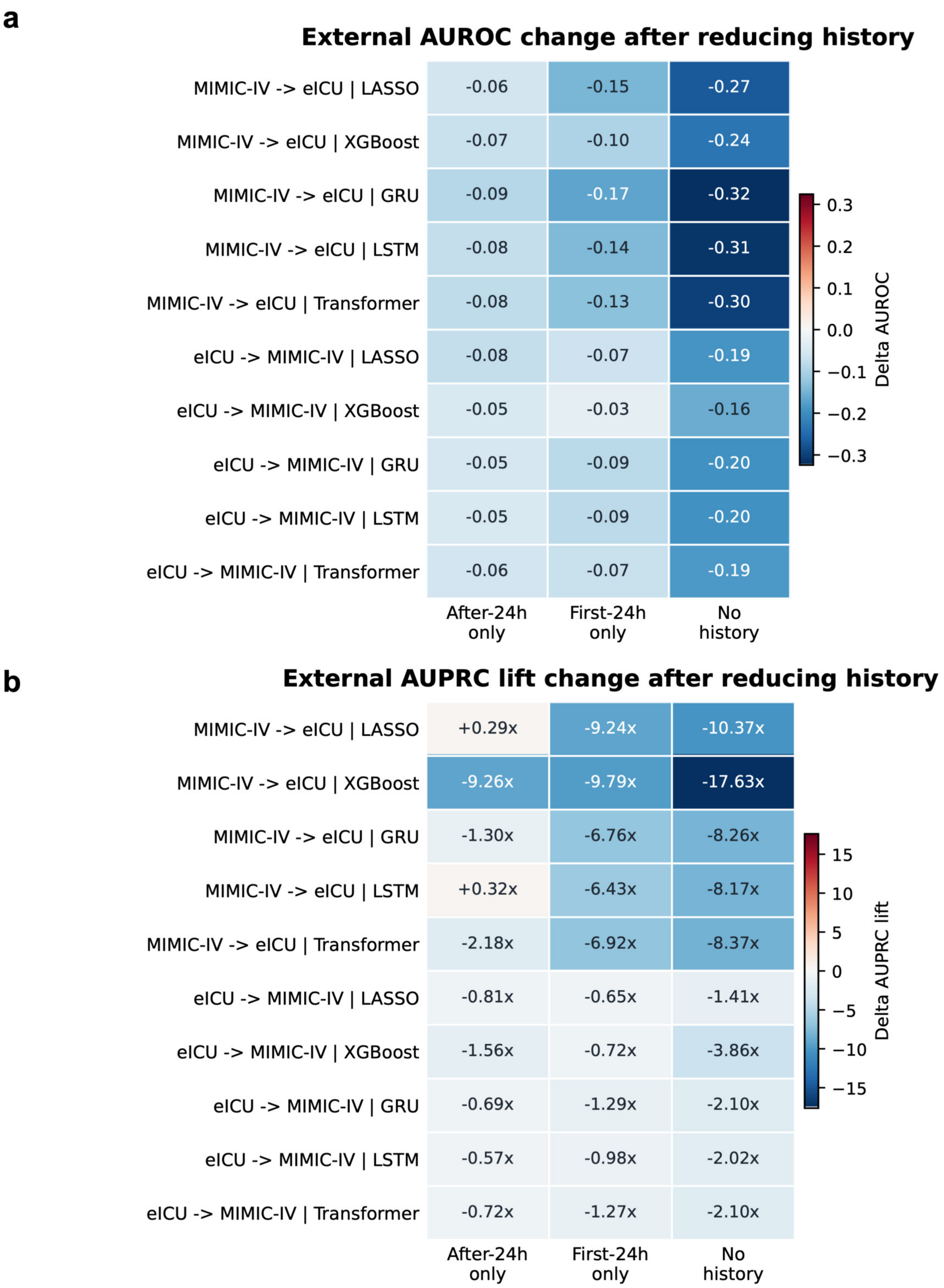


Figure S16: External performance change after reducing delirium-assessment history. Cells show each reduced-history variant minus `both_hist` for AUROC (**a**) and AUPRC lift (**b**) in each transfer direction and model. Negative values indicate lower performance than the both-history reference. AUPRC lift differences are arithmetic differences between lift values, expressed with the × suffix.

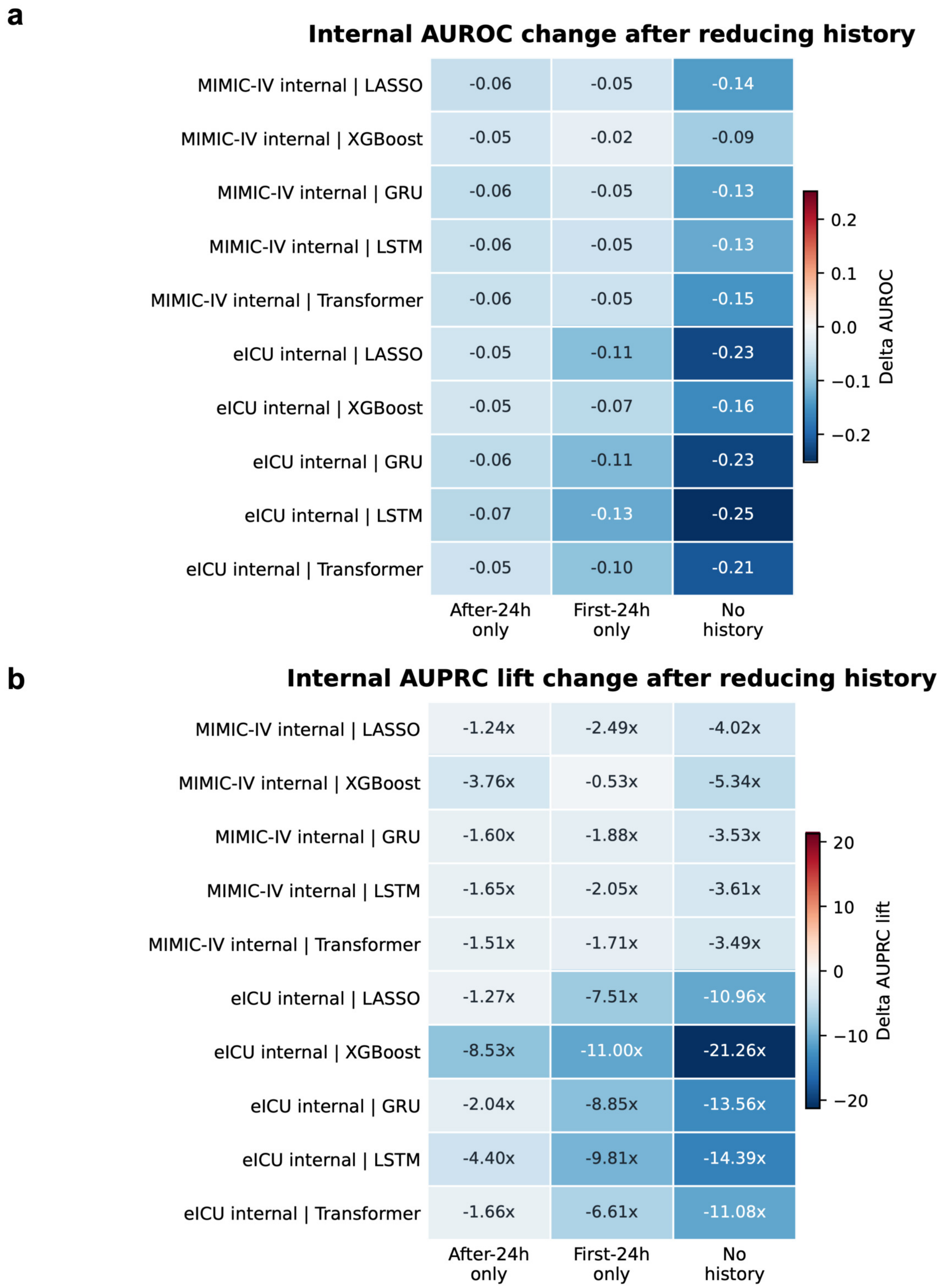


Figure S17: Internal performance change after reducing delirium-assessment history. Cells show each reduced-history variant minus both_hist for internal AUROC (**a**) and AUPRC lift (**b**) in MIMIC-IV and eICU. Negative values indicate lower performance than the both-history reference.

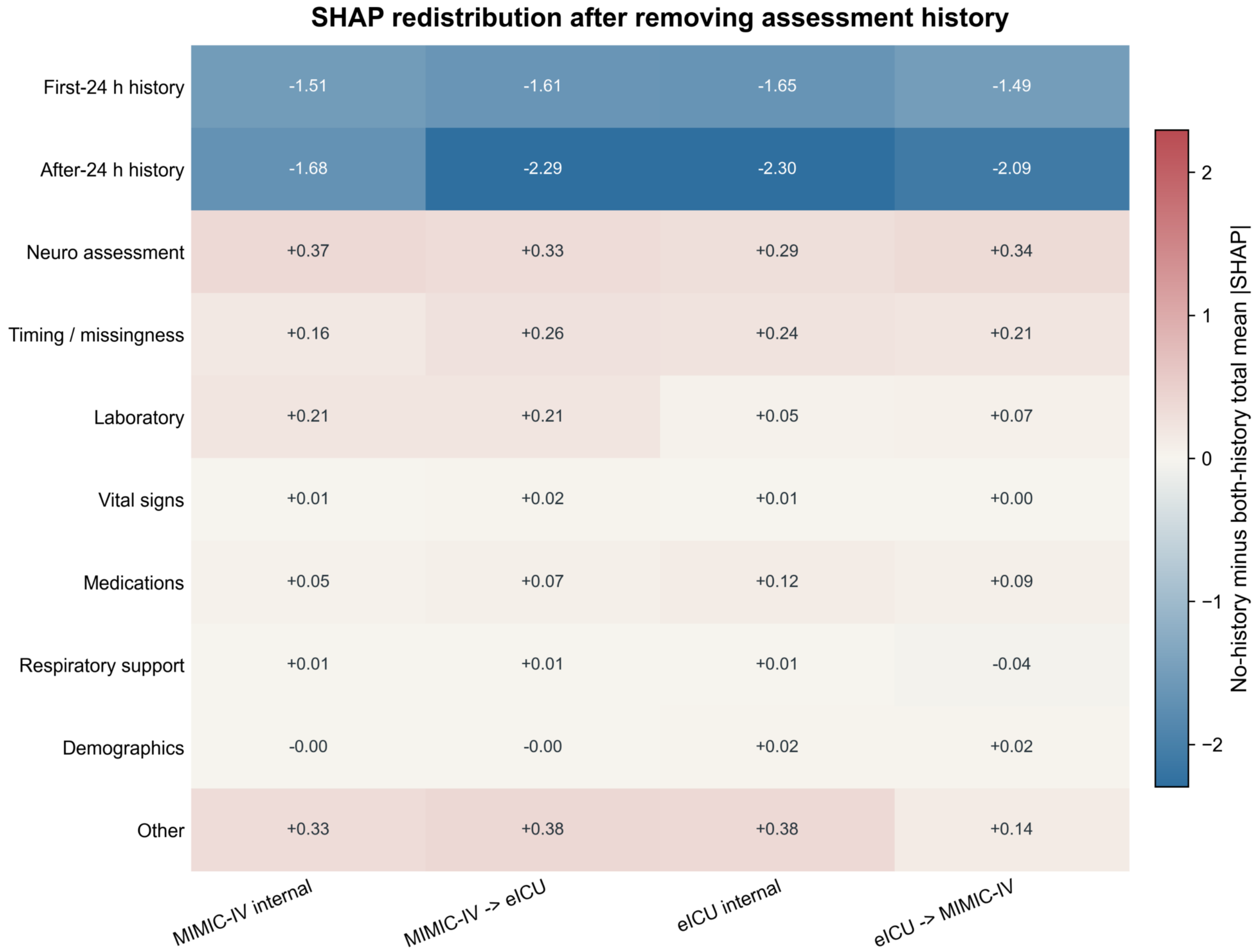


Figure S18: XGBoost attribution redistribution after removing assessment-history features. Cells show the no-history minus both-history difference in total mean absolute SHAP value for each feature domain and evaluation setting. Negative values in the first-24 h and after-24 h history rows are structural because those features were removed; positive values in retained domains indicate redistribution of model attribution. The comparison is within each refitted model and does not imply causal feature importance.

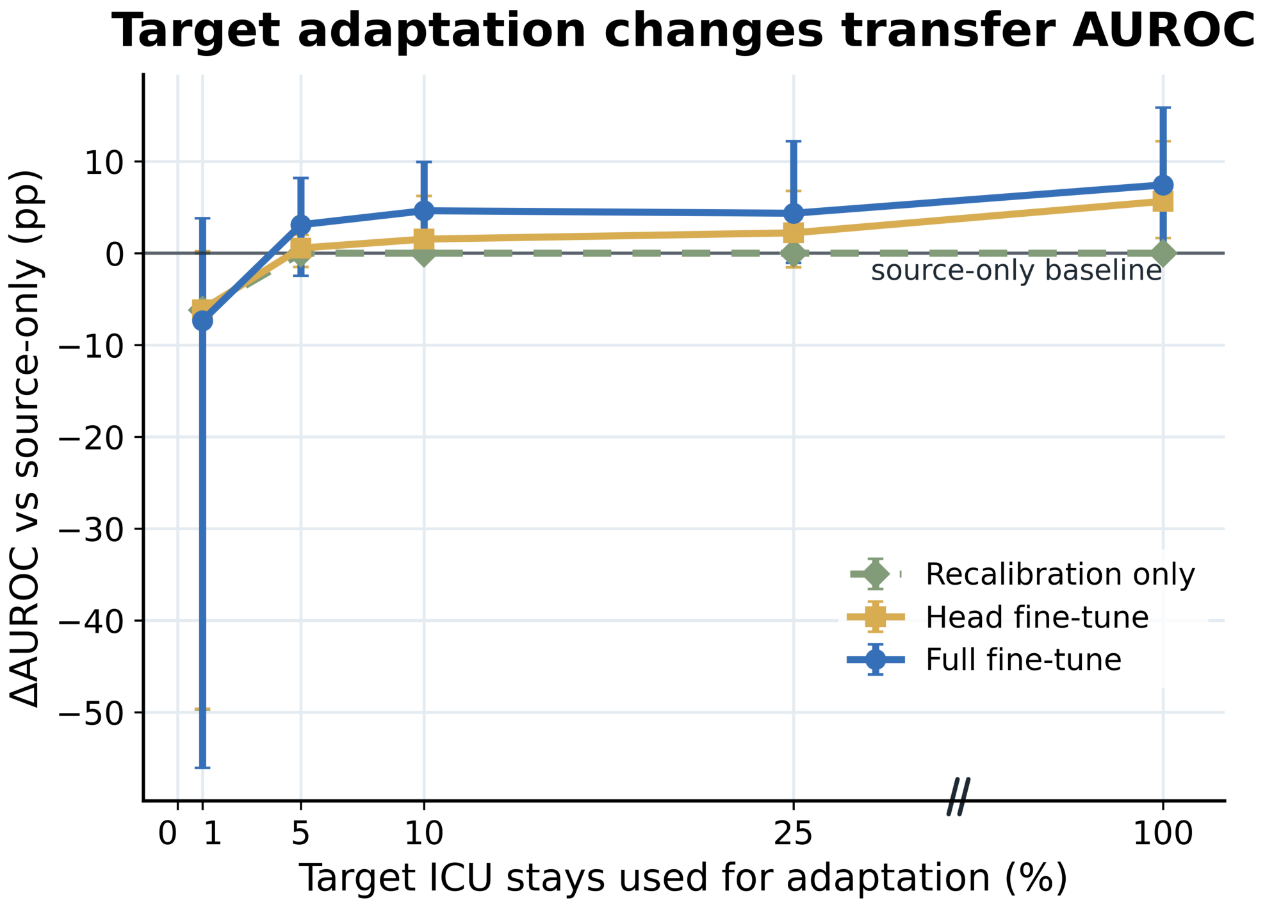


Figure S19: Change in external AUROC during supervised target-label adaptation. Points show mean within-run paired differences from the corresponding source-only GRU model for recalibration only, head-only fine-tuning and full fine-tuning. Target-development subsets were sampled by ICU stay, whereas AUROC was calculated over eligible prediction windows. Intervals show the observed minimum–maximum range across eight direction–run results (four model-training runs in each transfer direction), not confidence intervals.

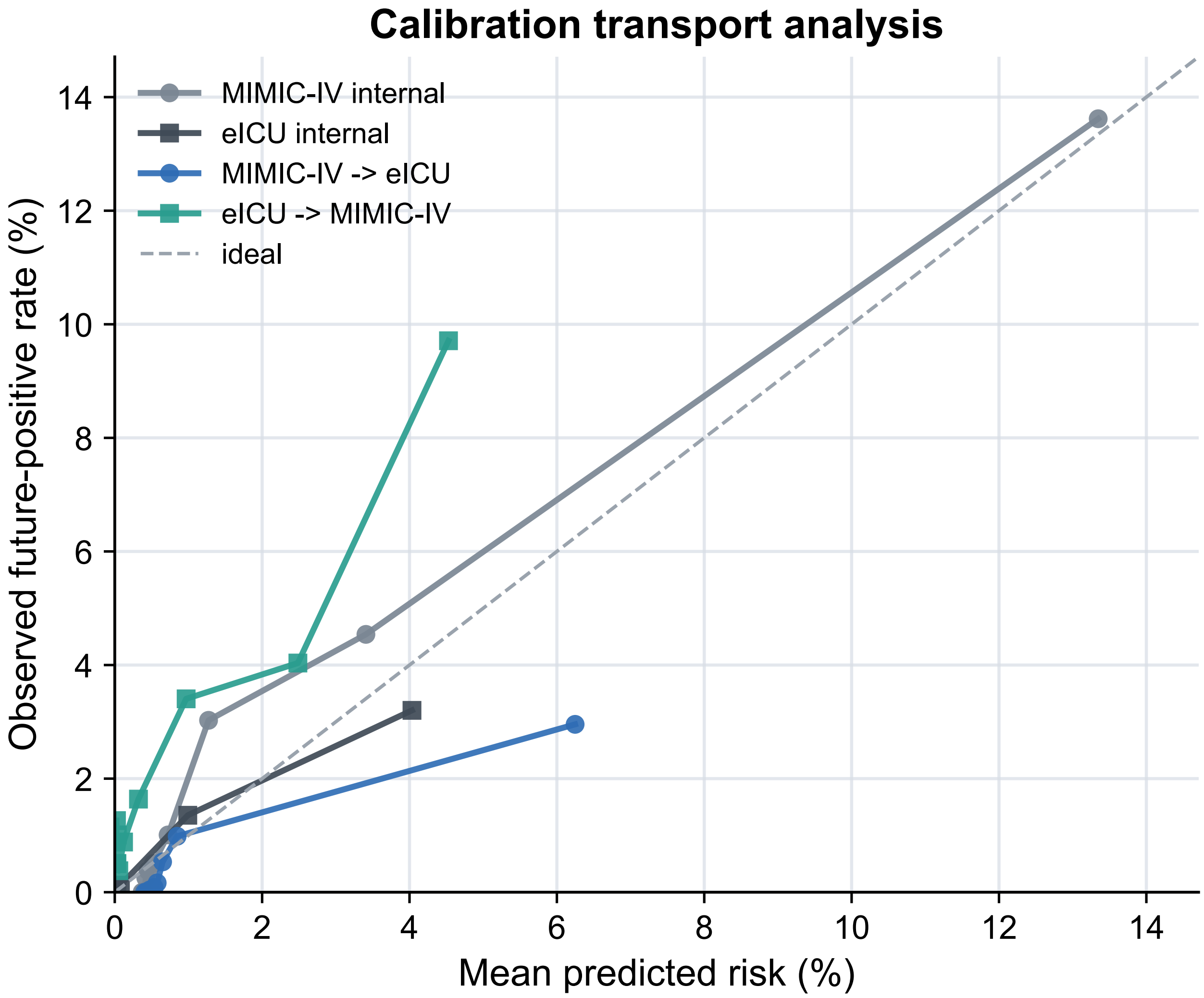


Figure S20: Calibration transport analysis. Calibration-bin observed future-positive rates are plotted against mean predicted risks for internal eICU and MIMIC-IV evaluation and for bidirectional source-only external validation. The diagonal denotes ideal calibration. Bins were constructed over eligible prediction windows and may contain multiple rows from the same ICU stay; the panel therefore evaluates probability transport at the window level and is not an ICU-stay policy analysis. Because events were sparse, binned patterns are descriptive and non-finite or non-positive calibration-slope estimates were treated as model-failure or instability flags rather than perfect calibration.

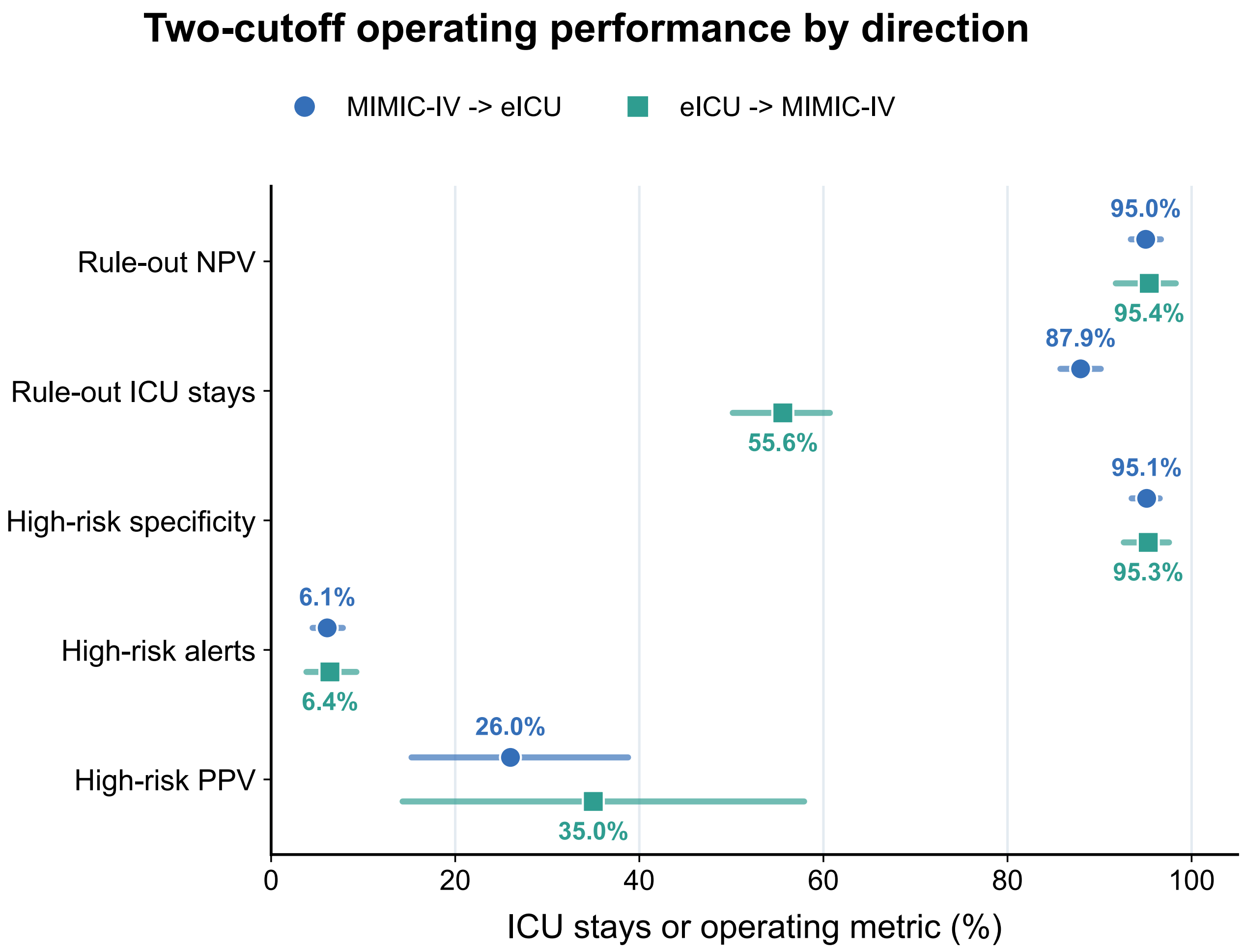


Figure S21: Exploratory same-test-selected direction-specific two-cutoff analysis. Low and high cutoffs were selected separately within the same external-transfer test ICU stays used for evaluation: 2.98% and 15.44% for MIMIC-IV-to-eICU transfer, and 1.39% and 5.00% for eICU-to-MIMIC-IV transfer. Points show operating estimates and intervals show percentile 95% intervals from 1,000 ICU-stay bootstrap resamples with selected cutoffs fixed; selection was not repeated within resamples. Because cutoff selection and evaluation used the same external test data, this analysis is descriptive and does not support the development-selected frozen-policy result in main Fig. 6e or any prospective actionability or benefit claim.

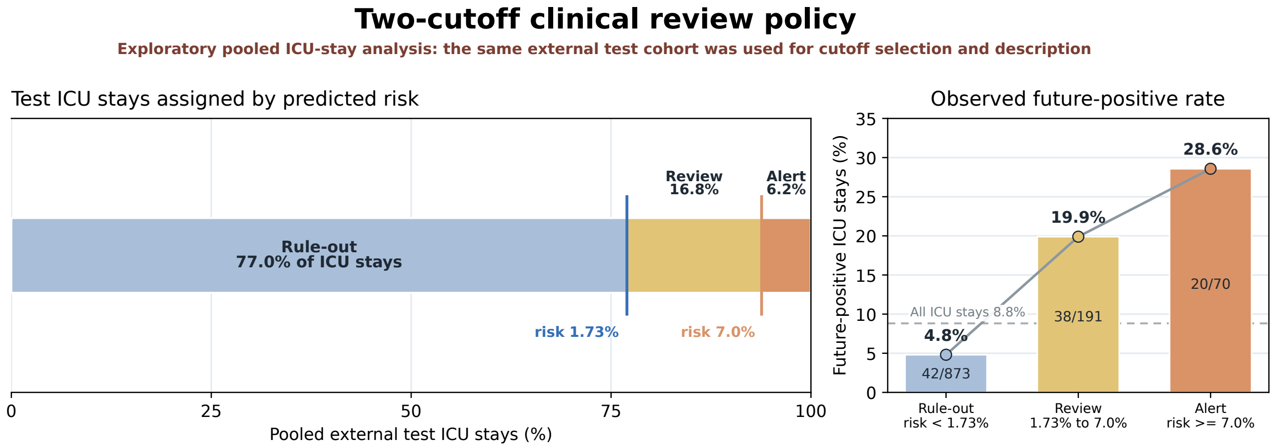


Figure S22: Exploratory pooled same-test-selected ICU-stay two-cutoff display. Each ICU stay was summarized by its maximum calibrated prediction-window risk. The pooled external-test cohort comprised 1,134 ICU stays; the same cohort was used to select the 1.73% and 7.00% cutoffs and to describe zone allocation and future-positive rates. The displayed operating estimates are therefore post hoc descriptions rather than independent policy validation. Main Fig. 6e instead uses development-selected cutoffs and external-test prediction-row denominators; deduplicated workload from the frozen policy is reported in Supplementary Table S13. This analysis does not support the frozen-policy result or any prospective actionability, intervention or benefit claim.

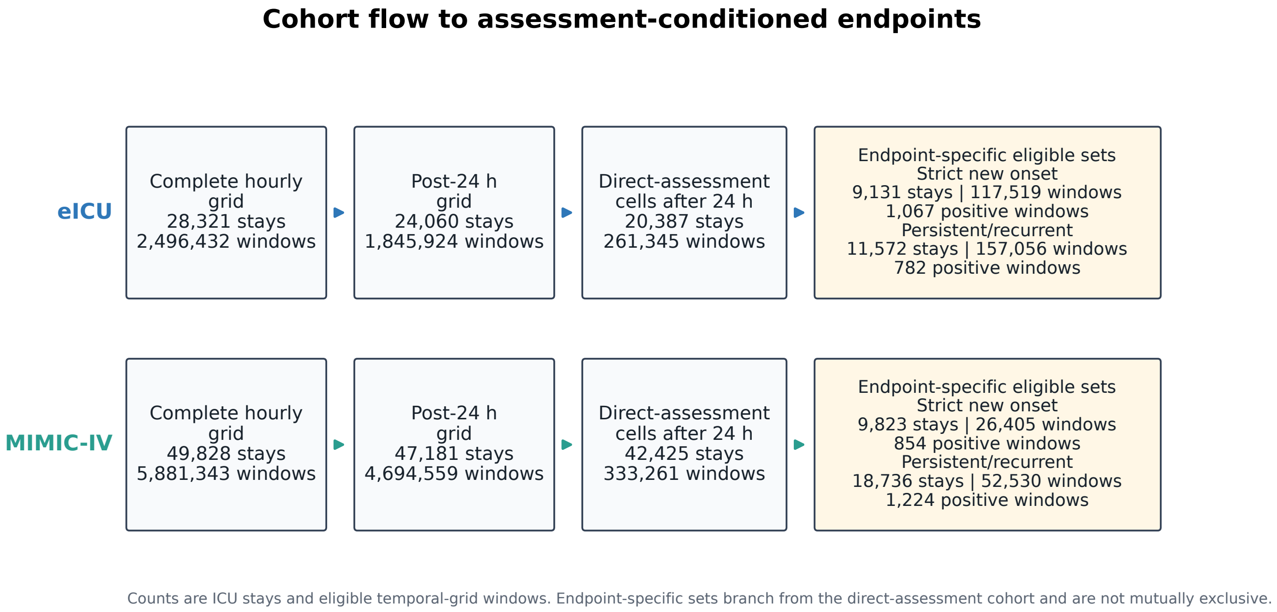


Figure S23: Cohort flow to the assessment-conditioned refined endpoints. Each lane shows the complete hourly grid, the post-24 h grid, direct-assessment cells after 24 h and the two endpoint-specific eligible sets. Counts are ICU stays, temporal-grid windows and endpoint-positive windows. Strict new-onset and persistent/recurrent sets branch from the direct-assessment cohort and are not mutually exclusive. No rows or counts were simulated.
